# CoAI: Cost-Aware Artificial Intelligence for Health Care

**DOI:** 10.1101/2021.01.19.21249356

**Authors:** Gabriel Erion, Joseph D. Janizek, Carly Hudelson, Richard B. Utarnachitt, Andrew M. McCoy, Michael R. Sayre, Nathan J. White, Su-In Lee

## Abstract

The recent emergence of accurate artificial intelligence (AI) models for disease diagnosis raises the possibility that AI-based clinical decision support could substantially lower the workload of healthcare providers. However, for this to occur, the input data to an AI predictive model, i.e., the patient’s features, must themselves be low-cost, that is, efficient, inexpensive, or low-effort to acquire. When time or financial resources for gathering data are limited, as in emergency or critical care medicine, modern high-accuracy AI models that use thousands of patient features are likely impractical. To address this problem, we developed the CoAI (<u>Co</u>st-aware <u>AI</u>) framework to enable any kind of AI predictive model (e.g., deep neural networks, tree ensemble models, etc.) to make accurate predictions given a small number of low-cost features. We show that CoAI dramatically reduces the cost of predicting prehospital acute traumatic coagulopathy, intensive care mortality, and outpatient mortality relative to existing risk scores, while improving prediction accuracy. It also outperforms existing state-of-the-art cost-sensitive prediction approaches in terms of predictive performance, model cost, and training time. Extrapolating these results to all trauma patients in the United States shows that, at a fixed false positive rate, CoAI could alert providers of tens of thousands more dangerous events than other risk scores while reducing providers’ data-gathering time by about 90 percent, leading to a savings of 200,000 cumulative hours per year across all providers. We extrapolate similar increases in clinical utility for CoAI in intensive care. These benefits stem from several unique strengths: First, CoAI uses axiomatic feature attribution methods that enable precise estimation of feature importance. Second, CoAI is model-agnostic, allowing users to choose the predictive model that performs the best for the prediction task and data at hand. Finally, unlike many existing methods, CoAI finds high-performance models within a given budget without any tuning of the cost-vs-performance tradeoff. We believe CoAI will dramatically improve patient care in the domains of medicine in which predictions need to be made with limited time and resources.

## 1 Main

Clinical risk prediction scores have a long history in medicine, and the number of such scores is rapidly increasing. The risk score database MDCalc.com hosted 80 clinical risk calculators in 2013 and over 500 in 2019, predicting adverse outcomes in conditions ranging from sore throat to heart failure [1]. In recent years, there has been an explosion of interest in using techniques from artificial intelligence (AI), including those developed in the subfield of machine learning (ML), to make clinical predictions. AI models use images of skin and eyes to classify cancer [2] and diabetic retinopathy [3], use waveform data such as electrocardiograms to classify heart arrhythmias [4], and use comprehensive medical record data to predict patient diagnoses and surgical emergencies [5, 6]. These models promise to make clinical outcome prediction easier and faster for healthcare providers; this possibility is especially important in areas such as emergency medicine and critical care, where providers’ time and attention are at a premium.

However, both existing and AI-based clinical risk scores suffer from a common drawback; they generally assume that all of the *features* (i.e., clinical variables) in the training set are known at prediction time, though acquiring all of these features in order to diagnose an individual patient may be impractical. Even sparse models, which select and use only a small number of features, do not account for the time or effort required to acquire those features. In time- or resource-constrained fields such as emergency and critical care medicine, important features are often missing due to time and attention limitations. For example, from 1995 to 2009, Emergency Medical Service (EMS) providers in Washington State spent a median of just 16 minutes on the scene of trauma incidents [7]. The median time from EMS dispatch to arrival at the hospital was 48 minutes, just under the initial “golden hour” within which treatment affords the best chance of preventing death. These and other time-limited, resource-intensive healthcare situations leave little time for deploying feature-rich AI predictions. A useful alternative AI approach would be to account for real-world limitations by jointly optimizing for data gathering cost – e.g., time, effort, or money – as well as accuracy. Such a model could learn on massive datasets with many features and select the optimal subset for prediction within any time, effort, or monetary budget. Most importantly, it would preserve the high accuracy of AI models while turning the heuristic process of feature selection into a principled optimization problem that the model can automatically solve.

To train prediction models with a principled joint optimization, we present a new AI framework, named CoAI (<u>Co</u>st-aware <u>A</u>rtificial Intelligence; Figure 1). CoAI calculates each feature’s predictive power (Figure 1a) and uses expert annotations of feature cost (Figure 1b) to choose the optimal set of features for any cost budget (Figure 1c). Given a new patient and a feature budget for prediction, CoAI can recommend which features to gather and make accurate predictions of patient risk given those features. Its main benefits include the abilities: (1) to quantitatively optimize the tradeoff between prediction performance and feature cost, yielding accurate *and* low-cost predictions; (2) to make *any* predictive model (e.g., deep neural networks, gradient boosted trees, etc.) cost-sensitive, dramatically increasing user choice; and (3) to train the model significantly more efficiently and with less hyperparameter tuning than existing cost-sensitive prediction approaches. We have released CoAI as open-source software compatible with the popular Scikit-Learn API for training AI models [8] ^1^. We compare CoAI against existing clinical risk scores and recent AI methods for cost-sensitive prediction, including cost efficient gradient boosting (CEGB) and a reinforcement-learning (RL) based method called “classification with costly features” (CWCF) [9, 10, 11] on three clinical tasks (Figure 2; Method 1).

**Figure 1:**
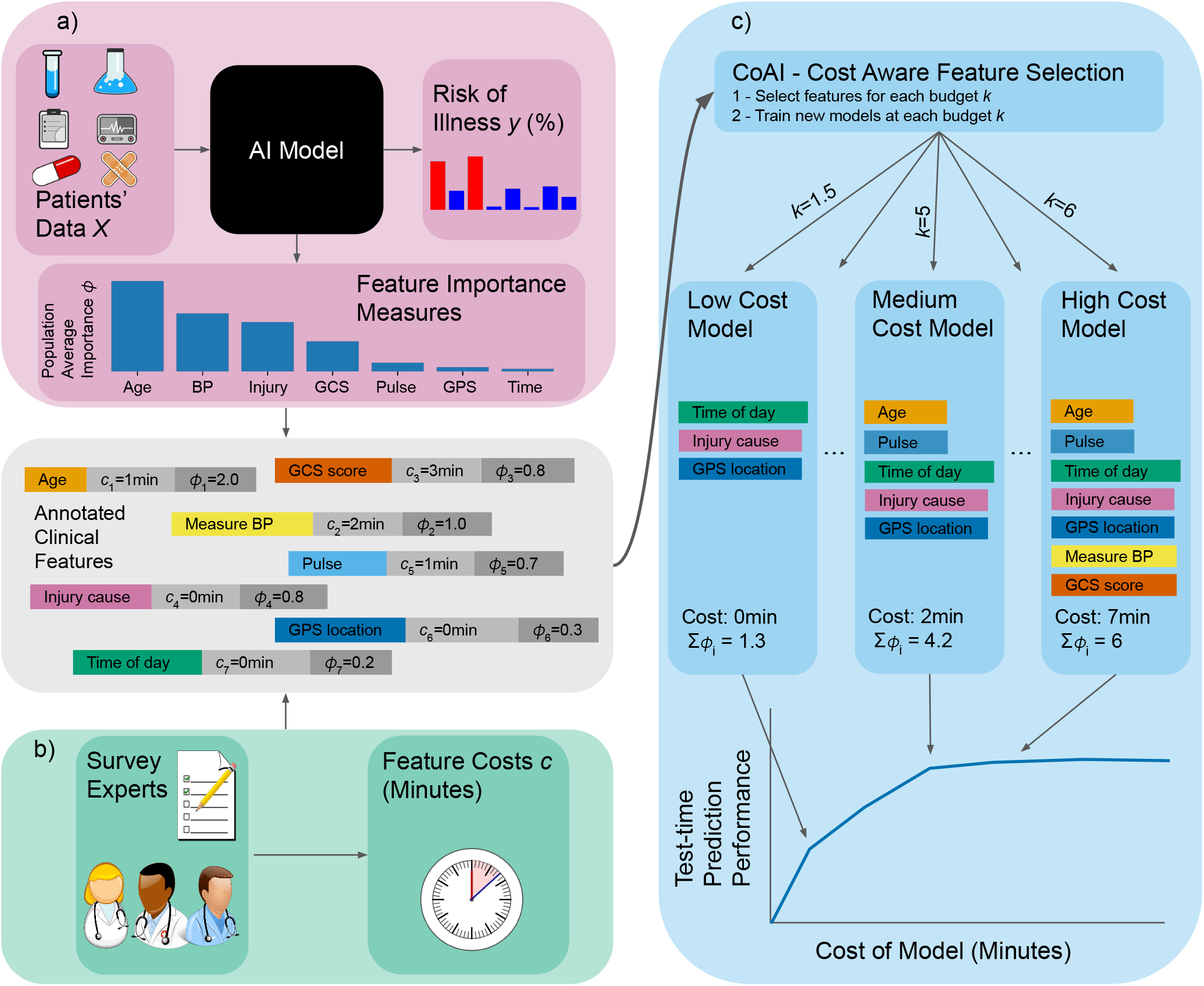
Overview of CoAI framework. Clinical features are annotated based on two different sources. (a) The *importance* of each feature is calculated by training an AI predictive model on the full data including all features and applying additive eature attribution methods to that model. The result is a single number summarizing the predictive power of each feature. By surveying clinical domain experts, the *cost* of gathering each feature is estimated. Costs considered in this paper nclude time cost (minutes) and financial cost (dollars), although it could be any numeric quantity. (c) (top) The CoAI algorithm takes as input all features, costs, and importance values and selects appropriate feature subsets for any cost budget. It trains a new AI model for each feature subset and cost budget that is desired. (bottom) Training CoAI models with multiple such budgets results in a cost-performance tradeoff curve, as larger budgets allow for gathering more eatures, which leads to higher-accuracy predictions.

**Figure 2:**
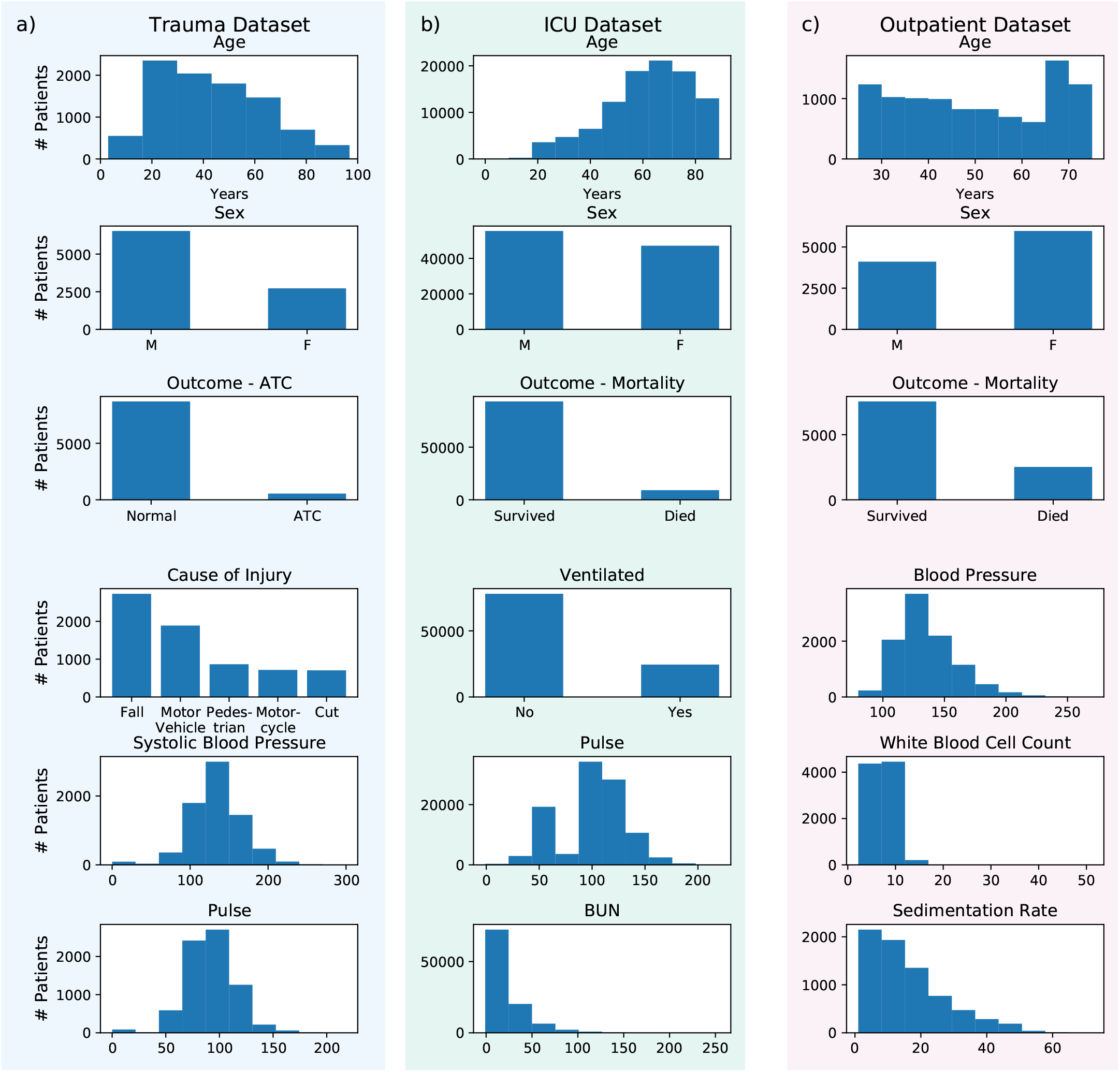
Histograms providing statistics for the (a) trauma, (b) ICU, and (c) outpatient datasets. The trauma and ICU datasets were used for benchmarking CoAI against existing clinical risk scores, while the trauma and outpatient datasets were used for benchmarking CoAI against existing AI methods for cost-sensitive prediction. The first three rows for each dataset show the distribution of age, sex, and outcome of interest for each dataset, respectively. The bottom three rows show the distribution of the next three most important features in each dataset, as measured by feature importance (Method 5). Notably, the trauma dataset and ICU dataset have clear age bias (younger patients are more likely to have traumatic injuries and older ones are more likely to end up in the ICU), while the outpatient dataset has a more uniform distribution as it was designed to be representative of American adults.

Our first cost-sensitive prediction task involves acute traumatic coagulopathy (ATC) in our “trauma dataset” – 14,000 emergency room visits and 45 features from the trauma registry of Harborview Medical Center, an urban Level-I trauma center (Figure 2a). ATC is an increased bleeding tendency involving anticoagulation and clot breakdown affecting up to 30% of severely-injured trauma patients [12]. ATC is associated with acute kidney and lung injury, increased transfusion needs, multiple organ failure and an 8-fold increased risk of early death [13, 14]. Trauma patients with ATC require complex care and rapid mobilization of hospital resources including massive blood transfusion protocols and surgical teams [15]. ATC diagnosis is currently based on coagulation testing in the hospital, which delays this time-critical diagnosis and complex healthcare response [16]. Therefore, our goal is to identify trauma patients at high risk of ATC as early as possible before arrival at the hospital to enable faster hospital-based life-saving interventions. Triage is only one of many tasks EMS providers must perform in trauma responses, so we aim to minimize the time required to gather input features. We selected prehospital features from the Harborview trauma registry, identified the data-gathering cost in minutes for each feature by surveying local experienced EMS providers (Figure 3d; Method 2; Supplementary Figure 1; and Supplement Sections 2-3), trained a CoAI ATC prediction model, and compared its cost and predictive performance with existing tools that predict ATC from prehospital data [17, 18]. One such model, the Prediction of Acute Coagulopathy in Trauma (PACT) score, required up to an estimated tenfold more data-gathering time than EMS providers reported being willing to spend during trauma responses, indicating the need for cost-aware modeling [17].

**Figure 3:**
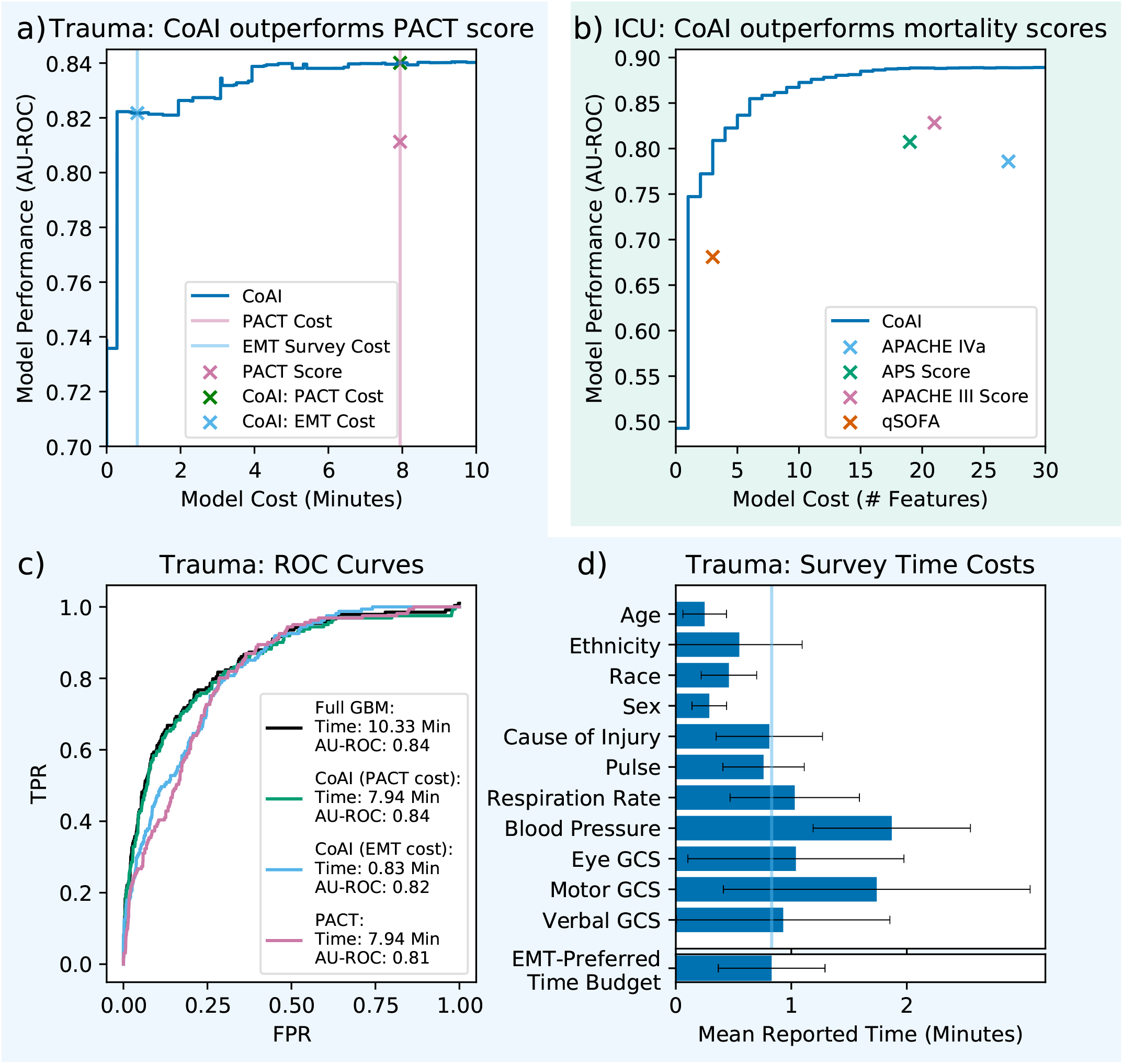
CoAI improves prediction performance and model cost over existing clinical models. a) For the trauma dataset, CoAI outperforms the existing PACT score. The right pink line is the measurement cost of the PACT model (7.94 minutes), and the left blue line is the EMS provider-preferred time budget (50 seconds). b) For the ICU dataset, CoAI outperforms four widely used clinical models across a range of potential cost budgets. c) Comparison of ROC plots at clinically important points on the cost-performance tradeoff curve for the trauma dataset. A gradient boosting machine trained on all features serves as a performance ceiling, requiring 10.33 minutes to gather necessary data. The PACT score requires 7.94 minutes and has worse AU-ROC. CoAI’s 7.94-minute model, however, can match the performance ceiling given by the full GBM. CoAI within the EMS provider-preferred time budget of 0.83 minutes still outperforms PACT. Measurement times for prehospital features gathered in a survey of EMS providers (except for features available at dispatch or indicating procedures performed, which were considered free). Normal 95 percent confidence intervals are shown as black lines. The survey also asked respondents to list what they believed was an appropriate amount of time to use a risk prediction tool (bottom bar, blue line; see Supplement Section 3).

We also examine the problem of in-hospital mortality prediction in critical care patients in our “ICU dataset” – 140,000 patient stays and 43 features from intensive care units (ICUs) across the United States (Figure 2b). ICU mortality is an important prediction target because it is (1) prevalent, with mortality rates in United States ICUs as high as 19 percent (2) costly, with ICU expenditures constituting 14 percent of hospital costs, and (3) highly variable across patients and hospitals even after adjusting for baseline patient characteristics [19]. For this task, we define cost simply as the *number of features* in the model. This is motivated by the fact that the mortality risk scores with the highest accuracy, such as the Acute Physiology and Chronic Health Evaluation (APACHE) model, are considered impractical for clinical use because they require a large number of features [20]. Models designed to be more efficient for clinical use, such as the Sequential Organ Failure Assessment (SOFA) and quick SOFA (qSOFA) scores, use as few as 3 features but suffer reduced performance as a result [21]. Our goal is to provide a single method that optimally trades off cost with performance and can provide accurate predictions using any number of features deemed clinically feasible.

Finally, we examine 10-year mortality prediction in an “outpatient dataset” from the long-running National Health and Nutrition Examination Survey (NHANES) – 13,000 outpatients and 35 features in outpatients across the United States (Figure 2c) [22]. Here, the goal is to minimize the *financial* cost in dollars of the data used by the model as estimated using Medicare fee-for-service data (Method 3; Supplementary Figure 2) while preserving prediction accuracy. This task uses a unique patient sample representative of the United States population. We choose such a dataset because mortality prediction is a ubiquitous clinical task; a national survey found that internal medicine physicians were asked to predict patient lifespan roughly once per month but felt ill-prepared to do so [23], and (2) a model that is applicable to all 480 million annual primary care visits in the United States [24] is an important case in which to lower the financial cost of risk scores. While long-term mortality scores have been developed for many specific diseases and patient subpopulations [25, 26, 27], we are not aware of a commonly-used outpatient mortality risk score applicable to the general primary-care population. We hypothesize this is due in part to the prohibitive expense of gathering data for routine mortality prediction, and hope to show that accurate, low-cost predictions can be made in this setting.

Across all three tasks, CoAI consistently improved predictive performance and lowered cost relative to both existing clinical risk scores and existing AI-based methods. CoAI bridges the gap between AI-based predictive models and the real-world constraints of clinical practice by ensuring that predictive models do not impose undue burden on their users. We believe this work will improve the accuracy of clinical risk predictions while ensuring that such predictions are made efficiently enough to have a real-world impact on patient care.

## 2 Results

### 2.1 CoAI Framework

Because of the wide array of prediction tasks and modeling strategies used in developing clinical risk scores, we developed the CoAI (Cost-aware AI) framework, which can be applied to any predictive model (called the *base model*) to make it cost-aware (Figure 1; Method 4). CoAI takes as input a training data set, consisting of patient features *X* and prediction labels *y* across patients, *costs c*_*i*_for measuring each feature *i*, and a *budget k* representing total acceptable cost for a predictive model. The goal of CoAI is to select a specific feature set *S*, with total cost no greater than *k*, that yields the best predictive performance given the budget.

This task is computationally challenging because, in general, the exact predictive value of a feature set is unknown without trying to train the model with that specific set of features. Previous approaches which are based on reinforcement learning (RL) attempt to directly search the exponentially large space of all possible feature sets, while others, such as decision tree-based approaches, simplify the problem via greedy search [9, 10]. The idea of CoAI is to find a feasible solution without enumerating all possible feature subsets and with a more optimal selection of features than greedy search approaches. This is enabled by calculating a single quantitative measure of predictive power for each feature, *ϕ*_*i*_and defining the predictive power of a feature *set S* as ∑_*i*∈*S*_*ϕ*_*i*_. We then select the feature set *S* that maximizes ∑_*i*∈*S*_ *ϕ*_*i*_ subject to ∑_*i*∈*S*_ *c*_*i*_ ≤ *k*, which is a knapsack problem that can be efficiently solved.

Our approach is motivated by the use of Shapley values for the feature importance measure *ϕ*_*i*_, which guarantee a set of desirable theoretical properties [28]. First, Shapley values are *additive* – that is, they sum to the model’s output – making ∑_*i*∈*S*_ *ϕ*_*i*_ a natural way to calculate the importance of a group of features. Second, they are *consistent*, which means features that are unambiguously more important are guaranteed to have a higher *ϕ*_*i*_. Finally, and most importantly, Shapley values can be calculated for any model, making CoAI a model-agnostic method and increasing user flexibility. However, Shapley values also have fast implementations for many popular model types, including deep models as well as the tree and linear models we use in this paper [28, 29, 30, 31]. We calculate *ϕ*_*i*_ by training an instance of the base model on all features in *X* and setting *ϕ*_*i*_ equal to the mean absolute feature importance across all samples.

Once importance *ϕ*_*i*_ based on Shapley values has been calculated for each feature, we use existing specialized knapsack solvers to select the optimal feature set [32]. We then train a new, final base model on this feature set. We demonstrated that knapsack solvers performed better than alternative approaches, such as greedy and recursive feature elimination approaches, to find the best feature set given *ϕ*_*i*_ and *c*_*i*_ (Method 6; Supplementary Figure 3).

### 2.2 CoAI improves cost and performance of clinical predictions

We evaluated CoAI against existing clinical models (Figure 3) by training it on the (a) trauma and (b) ICU datasets – those for which there are existing clinical risk scores for comparison – with gradient boosting machines (GBMs) as the base model (Method 7). We plotted its predictive performance (area under the ROC curve, AU-ROC; higher is better) across a range of measurement budgets, resulting in a cost-performance tradeoff curve. Existing clinical risk scores are shown on the same plots as points with a fixed model cost and performance. CoAI exhibits the strongest performance at any cost budget in both datasets.

To determine how CoAI could improve prediction of ATC, we compared it to the Prediction of Acute Coagulopathy of Trauma (PACT) score (Figure 3a; Method 8; Method 9) [17]. PACT is a multivariable logistic regression developed for prehospital ATC prediction. It uses the following prehospital features: patient age, presence of prehospital CPR, presence of prehospital intubation, injury mechanism, Glasgow Coma Score, and shock index (first prehospital pulse/systolic blood pressure). In our survey of EMS providers, total time cost incurred to obtain all PACT features was 8 minutes (Figure 3d).

We compared ROC plots of PACT to those of CoAI at several clinically important points along the cost-performance tradeoff curve (Figure 3c). For the same time cost as the PACT score (8 minutes), CoAI performs as well as a cost-unconstrained model (0.84 AU-ROC) and exceeds PACT score performance (0.81 AU-ROC). We also determined a realistic time budget using our survey of EMS providers (Figure 3d), who reported being willing to spend 50 seconds using a predictive risk tool on average. This tightly constrained budget is about tenfold less time than the PACT score requires, but the performance of CoAI within this budget (0.82 AU-ROC) still exceeded PACT’s performance. Importantly, CoAI’s prehospital prediction performance compares favorably to existing *post* -hospital admission ATC models; a previous study of AI models for ATC achieved AU-ROCs from 0.83 to 0.86 using vital signs, blood gas measurements, and lab values gathered *after* patients entered the hospital [33]. CoAI attains similar performance using only tightly time-constrained prehospital data.

We also used these results to estimate the impact of deploying CoAI risk scores nationwide (Method 11, Supplementary Figure 4). We expected that CoAI would both increase sensitivity to ATC and reduce the time cost required to make predictions. By extrapolating from total EMS trauma calls in the National Emergency Medical Services Information System database and ATC base rates in our data, we estimate that 120,000 EMS trauma patients in the United States have ATC per year [14, 34, 35]. Assuming that providers are willing to tolerate 4 false positive alerts for every true positive [36, 37], deploying CoAI at the PACT budget would provide early warning of 36,355 more ATC cases than PACT. Deploying CoAI at the EMS-preferred time budget of 50 seconds would provide warning of 14,602 more cases than PACT *and* save 200,000 total hours of data-gathering time across all providers (Method 11 and Supplementary Figure 4).

For the ICU dataset (Figure 3b), the APACHE IVa score is known for its high accuracy, however, is difficult for clinicians to use at the bedside because it requires 27 features to be gathered (Method 8; Method 9). Conversely, the qSOFA score uses only 3 features but is much less accurate. CoAI outperforms qSOFA using only a single feature (admission diagnosis, AU-ROC 0.75) and outperforms APACHE IVa using only 3 features (AU-ROC 0.81). CoAI also outperforms the related APACHE III and APS scores at much lower model cost (5 features). Nationwide, we estimate that 450,000 US ICU admissions end in death per year [19]. At a 3.5-to-1 ratio of false to true positives, qSOFA could use 3 features to provide warning of 182,653 deaths, while CoAI with 3 features correctly predicts 389,809 deaths – an increase of 207,156 (Method 11 and Supplementary Figure 4).

### 2.3 CoAI builds more flexible models that outperform existing cost-sensitive AI methods

We used the trauma and outpatient datasets to demonstrate how CoAI improves cost and performance over other AI methods. We focus on these two datasets because they have non-uniform feature costs, which provides a more rigorous methodological evaluation. Figure 4 shows cost-versus-performance plots on the (a) trauma and (b) outpatient datasets, with models retrained 100 times on different train/test splits of the data. In these experiments, we train CoAI with both GBMs and logistic regression models as the base predictors to take advantage of its flexibility and model-agnosticism (Method 7). We compare CoAI to: (1) cost-efficient gradient boosting (CEGB), a popular and effective method for making cost-sensitive predictions with decision trees [9], which applies a fixed per-feature penalty whenever the tree model splits on a given feature for the first time, and (2) a reinforcement learning method called classification with costly features (CWCF) [10, 11], which penalizes an agent for selecting costly features and rewards it for producing correct classifications (Method 10-Method 13). Overall, CoAI consistently outperforms other AI methods. The best CoAI model in each plot has a significantly higher mean AUC, averaged across all possible budgets, than all other models, with *p* < 10^−5^ and *t* > 5.75 by paired-samples *t*-test for all comparisons (Method 14).

**Figure 4:**
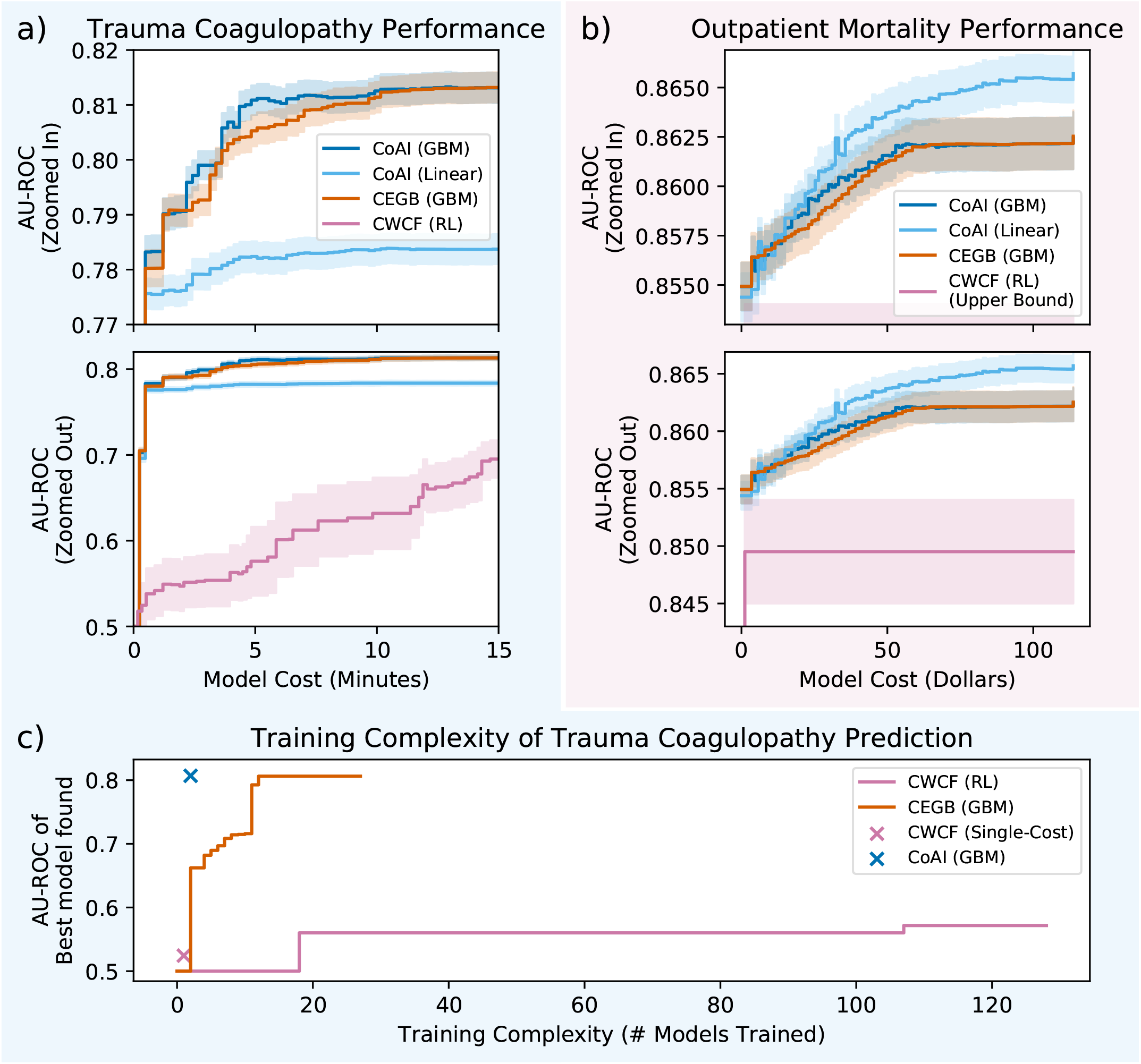
CoAI improves prediction performance, model cost, and training complexity over competitor methods. Lines are mean performance over 100 random train/test splits, and shaded bands are 95 percent normal confidence intervals. a) In the trauma dataset, linear models are not as effective as complex ones, but GBM-based CoAI has the highest performance at most budgets. CWCF performance increases with budget but never matches the other methods. b) In the outpatient mortality prediction task, both linear and GBM-based CoAI outperform CEGB and CWCF, and linear CoAI achieves the highest performance. c) Performance of best model found versus number of models trained for CoAI, CEGB, and CWCF. CoAI achieves the highest performance with only two model trainings.

For both datasets, cost-effective models reduce costs significantly without sacrificing performance, with CoAI and CEGB both plateauing in performance about halfway through the range of possible budgets. This implies that cost-aware learning can enable both trauma coagulopathy prediction in shorter periods of time (Figure 4a) and outpatient mortality prediction at lower cost (Figure 4b).

CWCF’s reinforcement learning (RL) approach also consistently underperforms other methods. Although RL has exhibited strong performance in binary classification [10, 11], it suffers in risk-stratification settings that use ROC and similar metrics because it produces hard classifications rather than a continuous *ranking* of patients by risk. We adapted CWCF to produce continuous outputs and ran fewer replicates to accommodate its slower runtime, but performance was still very low (Method 13).

The results in Figure 4 show several specific benefits of CoAI’s design relative to other cost-sensitive learning methods. First, CoAI is *model-agnostic*, i.e, it can be used with any predictive model. For the trauma dataset, CoAI with a GBM as the base model achieves the highest performance, likely because of nonlinear relationships in the data (Figure 4a). However, for the outpatient dataset, CoAI with a linear base model achieves the highest performance; in this case there is insufficient complexity in the data to warrant the additional overfitting that is possible with a GBM (Figure 4b). CoAI’s model-agnostic nature improves performance because it can use linear models when they are most appropriate and GBMs, or other complex models, when they are most appropriate.

Second, CoAI can easily use features that come in *groups* that each have a single acquisition cost. This situation is extremely common in AI tasks where features are redundantly encoded – such as nonlinear transformations of features or one-hot encoding – as well as in clinical medicine where many features (e.g., urine tests for blood and pH) can be acquired with a single lab test (urinalysis). CoAI naturally handles these situations because feature importance measures *ϕ*’s are additive and can be easily summed to yield *group* importances (Method 15). In the outpatient data, 27 exam findings and lab tests (groups) gave rise to 35 features, which expanded to 118 features after one-hot encoding. While each model could access all features, only CoAI could correctly place costs at a group level during training. While we adjusted the results of CEGB and CWCF at test time to account for grouped costs and calculated a generous upper bound for CWCF’s performance in this setting, CoAI still outperformed both methods, as shown on the cost-performance plot (Figure 4b, Method 13 and Method 14).

### 2.4 CoAI achieves orders-of-magnitude greater efficiency than other cost-sensitive AI models

Here, we demonstrate that CoAI also achieves better training complexity than other models. All cost-sensitive models require a tuning parameter to control the tradeoff between accuracy and cost. For CoAI, this parameter is the budget itself. Given a fixed target budget, CoAI always yields the optimal model within that budget in a constant number of training rounds. For CEGB, however, the tuning parameter is unitless (representing the ratio between model cost and loss in the optimization objective). Given a fixed target budget, this requires blind tuning until a good enough model is found that fits within the budget. CWCF supports both budget-based and unitless tuning parameters.

We tested the training complexity of all three models on a single train-test split of the trauma dataset, where we attempted to maximize performance at a single budget (the EMS provider-preferred time cost of 50 seconds) with each model (Figure 4c; Method 16). Blind tuning on CEGB with binary search requires training over 5 times more models than CoAI to reach a similar level of performance (13 total models). CWCF with unitless tuning takes a large number of trainings to yield even a small increase in performance and never reaches the same level of performance as CoAI or CEGB (128 total models). CWCF with a cost-based tuning parameter requires only a single model training, but yields very low performance. Only CoAI is able to offer high predictive performance with a low training complexity.

### 2.5 CoAI reveals high-yield features and dynamics of feature importance over time

We analyzed the order in which CoAI selected features to understand how it differs from other clinical risk scores. Figure 5 shows how feature rankings differed between CoAI and existing clinical models for the ICU and trauma datasets. For CoAI, the top features are listed in the order in which they were first added to the model as the budget was gradually increased. For each clinical risk score, the top features are listed in order of importance; for APACHE, importance is measured by loss reduction (Method 9) and for PACT, importance is measured by standardized regression coefficient. The qSOFA score weights all variables equally.

**Figure 5:**
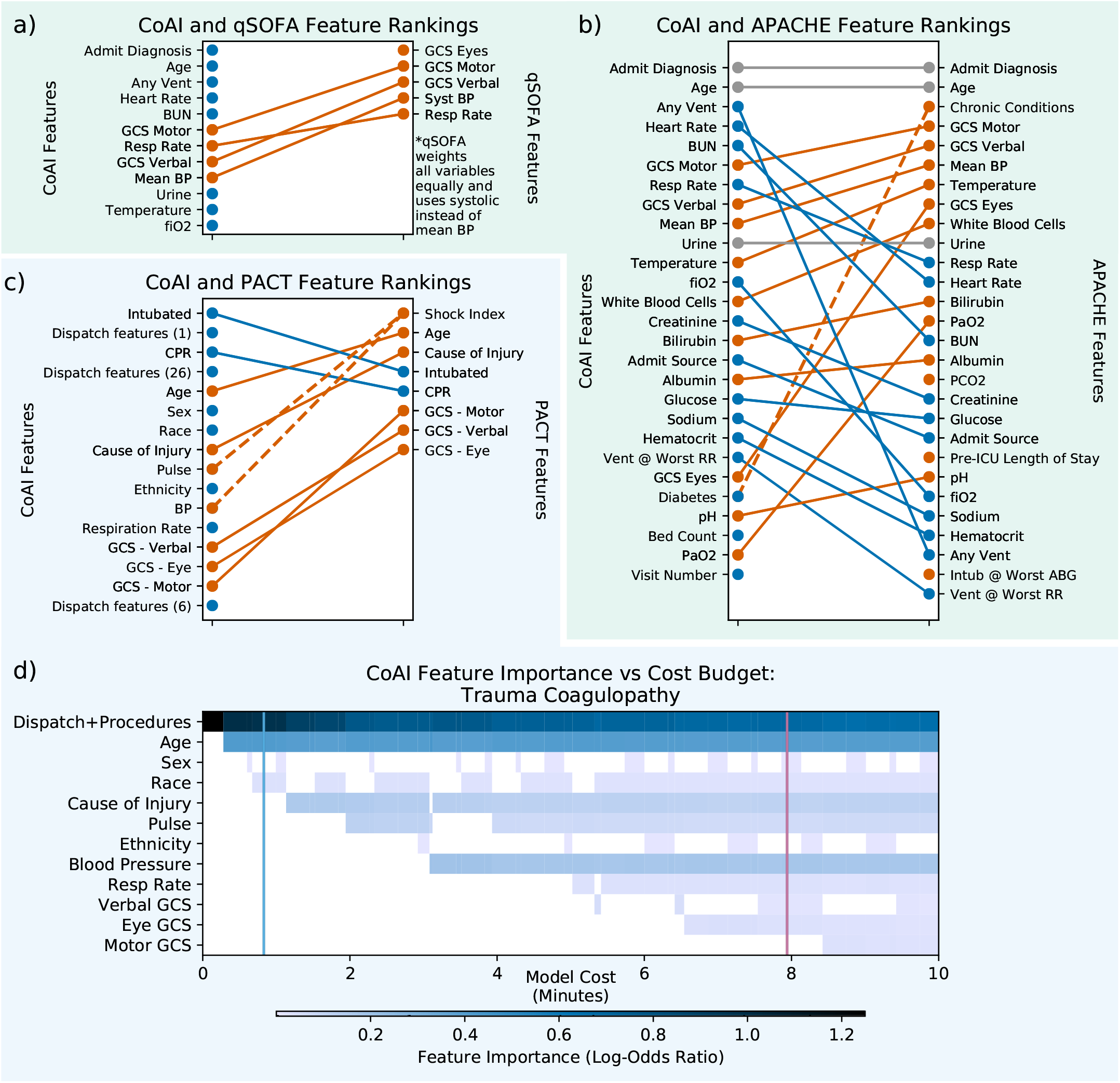
Importance of features selected by CoAI and other risk scores, ranked by order added to the model for CoAI and by regression coefficient or loss reduction for others (see Section 2.5). Orange features are ranked lower in CoAI than the corresponding clinical risk score, and blue ranked higher. Gray indicates no change. CoAI is compared to (a) qSOFA (b) and APACHE for the ICU dataset. It is compared to PACT (c) for the trauma dataset. All features are shown for qSOFA, APACHE, and PACT. For CoAI, the full feature list is shown in (c); in (b) 27 features are used to match APACHE and in (a) only 12 features are used for clarity (features are identical to (b)). Dashed orange line in (b) indicates that APACHE had access to several chronic conditions, while CoAI had access to only one – diabetes. Dashed line in (c) ndicates that shock index is calculated from both pulse and BP. (d) Each heatmap column shows the importance *ϕ*_*i*_ of each feature for a CoAI model trained at a particular budget *k* in the trauma dataset. Dispatch and procedure features have zero cost and are grouped into one row. Darker blues indicate more importance. Columns have varying widths since they are scaled to align with each model’s feature cost on the x-axis (in minutes). The left vertical blue line shows the EMS provider-preferred time budget; the right pink line shows the PACT time cost.

For the ICU data, CoAI and existing clinical models rely on different subsets of features (Figures 5a and 5b). Although CoAI and APACHE both rely on admission diagnosis and age, CoAI ranks ventilation, FiO2, heart rate, and blood urea nitrogen higher than APACHE, while ranking chronic conditions, PaO2 and the eye component of the Glasgow Coma Scale lower. The qSOFA model uses a small number of features, most of which are also used by CoAI, although CoAI also relies on many features not chosen by qSOFA. Notably, the higher-ranked CoAI features tend to be baseline information – age, diagnosis, and ventilation status – rather than specific vital signs. A similar situation arises with the PACT score in the trauma dataset; many PACT features are also important in the CoAI model, but CoAI prioritizes inexpensive data available at the time of dispatch before relying on the many vital signs used in PACT (Figure 5c). In particular, the intubation and CPR procedures are ranked more highly by CoAI than by PACT.

A unique aspect of CoAI is the opportunity to examine how changing budgets might affect the value of individual features. To investigate this, we examined the effects of changing time budgets on feature importance for the trauma CoAI models. The heatmap in Figure 5d displays the importance of each feature as a function of model cost as the budget *k* increases. This analysis reveals complicated dynamics of important features over time. For example, when very little time is available, dispatch and procedure information dominate the prediction (top row). Dispatch features include the patient’s geographical location and level of response e.g., Advanced Life Support or Basic Life Support, and procedures include CPR and intubation. This result agrees with recent data suggesting that prehospital procedures have high predictive value for predicting the need for hospital interventions such as massive transfusion [38]. However, when time budgets increase other features, such as vital signs, gain more significance and the importance of the earlier demographic features tends to decline. Occasionally, features like pulse are removed from the model entirely. Further, a model that adds features in a fixed order (e.g., greedy and recursive feature elimination methods, see Method 6) cannot make these sophisticated time and information tradeoff choices and would lose performance as a result. These results show that CoAI’s ability to train models within any budget is valuable not only for flexibility in deployment, but also as a way to better understand the predictive value of each patient feature. Further analysis of feature importance, as well as importance heatmaps for other datasets, are shown in Supplementary Figures 5-9.

## 3 Discussion

As AI and ML models become increasingly prevalent in healthcare, they risk imposing a large data-gathering burden on health care providers unless steps are taken to ensure such models can automatically select highly informative, easily acquired features. Our study is the first, to our knowledge, to survey clinical experts, build improved cost-aware clinical risk scores, and evaluate cost-aware models at operating points chosen by clinical providers.

Our framework, CoAI, is simple, flexible, and can efficiently adapt any predictive model to make cost-sensitive predictions. In the trauma dataset, CoAI’s sophisticated AI models and its automated choice of low-cost, high-information features produce predictions more accurate than existing clinical scores at less than one-tenth the data-gathering cost. Analogously, for ICU mortality prediction, CoAI outperforms existing risk scores using one-ninth of the total number of features. CoAI also makes accurate predictions at low monetary cost in an outpatient mortality prediction dataset, outperforming other AI methods for cost-sensitive prediction. Extrapolating these results across all trauma and ICU patients in the United States indicates the potential for CoAI-based risk scores to provide advance warning of tens of thousands of ATC cases and hundreds of thousands of deaths while providing substantial time savings – hundreds of thousands of hours of total provider time in the trauma dataset.

CoAI has several desirable properties that make it better suited to cost-aware clinical risk scoring than other AI methods. Its model-agnostic nature enables accurate, low-cost predictions in a wide variety of settings by using complex models for large or nonlinear datasets and simple models for small datasets or ones where linear relationships are expected. Its ability to handle grouped features also makes it a natural fit for data with feature transforms or one-hot encoding and for data where properties of the data acquisition process, such as lab tests that return multiple measurements, result in naturally grouped features.

CoAI demonstrates several other benefits relative to existing methods. Most existing tools do not make hard guarantees or provide worst-case bounds on feature acquisition cost – decision tree and RL methods may ask for features indefinitely so long as it improves prediction accuracy enough. CoAI imposes a hard time cutoff in its optimization so that users can be sure it will not recommend excessively costly features. This hard cutoff also serves as the cost-versus-performance tuning parameter, helping users to quickly acquire the best possible model for their desired budget without blindly tuning tens or hundreds of models as other methods do. While CWCF is also capable of using a hard cutoff tuning parameter, it performs hard classification and does not output probabilities of each class by default. This makes it difficult to use for risk stratification and significantly reduces performance on clinically important metrics like AU-ROC. Finally, CoAI always requests the same features within a given time budget, which increases predictability for healthcare providers; methods like RL or decision trees may ask for different features for different patients.

There are several avenues for future work with CoAI. Like other low-cost AI methods, CoAI assumes the cost of a feature set is additive – equal to the sum of individual feature costs. This may be reasonable in ambulances or outpatient clinics, where a single provider must perform tasks sequentially, but the additivity assumption may need to be relaxed in other settings, like large hospitals, where multiple providers can perform different exams and tests simultaneously. CoAI also does not fully account for feature interactions in which including a particular feature changes the relative value of other features. While CoAI performed better in our experiments than approaches like CWCF that explicitly account for feature interactions, a version of CoAI that accounts for interactions could improve performance even further.

Overall, we believe CoAI has demonstrated the potential to significantly improve clinical risk prediction. Its design treats the ease of gathering features just as importantly as the accuracy of predictions made using them. Our software is easy-to-use and integrated with existing open-source frameworks. We believe CoAI will make clinical risk scores more cost-sensitive, more accurate, and more effective at saving lives.

## Data Availability

Of our three datasets, the ICU and outpatient datasets are publicly available. The ICU dataset is available from the MIT eICU Collaborative Research Database but requires approval before download. The outpatient dataset is a subset of the NHANES I study. It is also uploaded to our Github repository along with our code. The trauma dataset is not publicly available due to patient privacy concerns.

https://eicu-crd.mit.edu/gettingstarted/overview/

https://github.com/suinleelab/coai

## Acknowledgements

This work was funded by the National Science Foundation [CAREER DBI-1552309, and DBI-1759487], American Cancer Society [127332-RSG-15-097-01-TBG], and National Institutes of Health [F30 HL 151074, R35 GM 128638, and R01 NIA AG 061132]. Thanks to Scott Lundberg, Ian Covert, and the Lee Lab for helpful discussions about this paper.

## Methods

### Method 1 Datasets

#### Trauma data

The trauma data used in this study were gathered over a 10-year period (2007 to 2017) and encompassed over 14,463 emergency department admissions for traumatic injury at a Level 1 Trauma Center. We selected 45 variables that were available in the pre-hospital setting, including dispatch information (injury date, time, cause, and location), demographic information (age, sex), and prehospital vital signs (blood pressure, heart rate, respiratory rate).

The outcome in this data was acute traumatic coagulopathy (ATC). We followed [17] and defined ATC as a binary outcome based on emergency department lab measurements of International Normalized Ratio (INR), where measurements greater than 1.5 were defined as coagulopathy.

#### ICU data

The ICU data were gathered from the public PhysioNet eICU repository [39]. The data come from over 142,139 patient admissions at 208 US hospitals between 2014 and 2015. While many features are available in this data, we selected 43 variables that had been preprocessed into a tabular format. Many of these variables are also used in the calculation of other risk scores, e.g., APACHE and APS. The outcome in this data was in-hospital mortality.

#### Outpatient data

The outpatient data were gathered from the NHANES I study, which is publicly available, with a reprocessed version for AI and ML recently released [22, 29]. The study gathered data on 13,442 individuals from 1971 to 1974, then followed up in 1992 to record 10-year mortality data. The NHANES data is unique among our datasets because: (1) it contains relatively healthy outpatients rather than relatively sick inpatients, (2) each row is an individual patient, as opposed to the trauma and outpatient dataset where stays are unique but patients may not be, and (3) it is a curated cross-sectional study rather than a convenience sample of patients who presented to the hospital, reducing dataset bias. We selected 35 features from this dataset including demographics, physical exam findings, and lab values. The outcome in this dataset was 10-year mortality.

#### Repeated measurements

In the trauma and ICU datasets, each sample is a patient visit, so a given patient who has visited the hospital multiple times could be represented as multiple samples in the dataset. In the outpatient dataset, each sample is a unique patient.

#### Data Processing

We standardized all variables to have 0 mean and unit variance. We mean-imputed missing data for input to all AI models except for CoAI in Section 2.2 because GBMs were the only AI model used in this section and could handle unprocessed missing data. Missing data in clinical models is discussed in Method 9. We treated categorical variables as categorical in LightGBM [40]. For the trauma and outpatient datasets, we used a random 64/16/20 train/validation/test split. For the ICU dataset, patients were grouped into four geographic regions of the United States; we split off 1 region as a test set and split patients from the other 3 regions into train/validation sets using an 80/20 split. Labels were binary for all datasets, and we included only the patients from each dataset for whom label data were available.

### Method 2 Prehospital Time Costs: Survey Methodology

We gathered time costs for the trauma dataset by surveying professional prehospital care providers from the Pacific Northwest. We designed a Qualtrics survey to gather information on previous EMS experience, past experiences with and thoughts about computerized risk scores, costs for each feature in terms of objective time and subjective effort, and overall impressions of the value of risk scores in prehospital medicine. The full list of survey questions, and summary data on responses, is in Supplement Section 2. Free text answers are not included to preserve anonymity. We used anonymous links to send the surveys to all employees of 3 major emergency medical services and collected all results from September 19 to October 25, 2019 for analysis. Costs for each feature were determined by the mean cost across respondents. The survey did not include questions on time or effort cost for variables already present at dispatch, for which a cost of 0 was assigned. While we gathered data on how long each procedure in our dataset took to perform, we set all procedure feature costs to zero, since entering procedure information in the risk score requires simply knowing whether the procedure was done.

### Method 3 Outpatient Monetary Costs

We assigned costs to features in the outpatient data by referencing Medicare data on payments for lab tests [41]. Physical exam and other free measurements were assigned a cost of zero. Unavailable costs were mean-imputed. The full list of feature costs is in Supplementary Figure 2.

### Method 4 CoAI Method

CoAI is a feature importance-based method for cost-sensitive prediction. Given a model *f* trained on the full dataset and a single patient’s features *x*_*j*_, CoAI uses recently developed axiomatic feature importance methods to assign an importance *ϕ*_*ij*_to each feature *i* such that the sum of all feature importances equals the risk prediction: ∑_*i*_ *ϕ*_*ij*_ = *f* (*x*_*j*_) [28]. These methods obey axioms guaranteeing heavier weighting of features with greater effects on the model’s output. By summing the absolute values of these importances across all data points, we can get a global measure of importance corresponding to the amount each feature impacted the model’s output across all predictions: *ϕ*_*i*_=∑_*j*_ |*ϕ*_*ij*_|.

CoAI uses these importance values to form an optimization problem that finds the best subset of features *F* – the one with the greatest sum of feature importances *ϕ*_*i*_– that do not exceed the cost budget. Mathematically, this can be written *F* = arg maxS *∑*_*i*∈*S*_ *ϕ*_*i*_s.t. ∑_*i*∈*S*_ *c*_*i*_ ≤ *k*, where *c*_*i*_is the cost of feature *i* and *k* is the cost budget. We can solve this well-known optimization problem, known as a *knapsack problem*, with arbitrarily small error in polynomial time with respect to the number of features due to a fully polynomial time approximation scheme (we use the Google OR-tools solver) [42, 32].

For any AI/ML model, this approach lets us find the exact optimal set of features that most impact the model’s prediction within any cost budget under the assumption that features’ effects are independent. In practice, the method works very well even when there are feature interactions. We also use a heuristic that substantially boosted performance in both our work and previous studies: after finding the best feature subsets for every budget, we retrain a new model using only those features [43]. For consistency, the new model is of the same model class with the same hyperparameters as the all-features model used to calculate feature importance. We tested several other methods for using feature attributions and costs to select the best feature subset within a cost budget, including a greedy algorithm, recursive feature elimination, and knapsack method without retraining. These methods did not match the performance of our knapsack method with retraining but may prove useful in specific situations (Supplementary Figure 3 and Method 6) Our approach is model-agnostic. Because general-purpose tools like LIME and SHAP can produce feature attributions for any AI/ML model, CoAI can also produce cost-effective versions of any such model. CoAI works best for model types that support fast calculations of axiomatic feature importances, such as linear, deep, and tree-based models. For all experiments in this paper, we use gradient boosted tree models (GBMs) or linear models for predictions and the SHAP package [29] to provide explanations (Method 5, and Method 7). We developed an implementation of CoAI that can build cost-sensitive versions of any model that supports the Scikit-Learn framework [8] and have released it as open-source software.

For interpretability purposes, we add small pseudocosts to zero-cost features (small enough that the sum of all such pseudocosts is less than the difference between any two non-zero-cost features). This is not strictly necessary, but does allow ranking of zero-cost features if desired.

### Method 5 Feature Attribution Methods

We needed measures of feature importance as well as feature cost to perform our CoAI analysis. We calculated feature importance using the SHAP (Shapley Additive Explanations) framework [28, 29], in which a feature’s importance is calculated with respect to a predictive model. The change of the model’s output when the feature is masked is recorded across all possible subsets of features, yielding an average change in prediction resulting from the inclusion of a feature in the model:

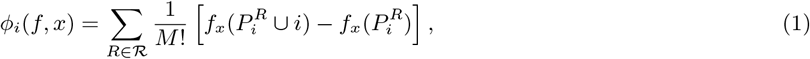

where *ϕ*_*i*_ is the importance of feature *i* in model *f* for data point *x*, ℛ is the set of all feature permutations, 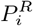 is the set of all features before *i* in the ordering *R, M* is the number of input features, and *f*_*x*_ is an estimate of the conditional expectation of the model output: *f*_*x*_(*S*) ≈ *E*[*f* (*x*) | *x*_*S*_] where *x*_*S*_ is the set of observed features.

SHAP estimates of feature importance can be calculated for any AI/ML model. Because we use GBM and linear models, we make use of fast algorithms for calculating SHAP values in decision trees and linear models [28, 29]. This improves runtime and reduces variability in importance estimates. It is also worth noting that CoAI is compatible with *any* feature attribution method that assigns a measure of importance *ϕ*_*i*_ to each feature – SHAP is not the only method of this type, but is model-agnostic, satisfies desirable axioms, and has fast specialized implementations for the models we consider here.

### Method 6 Alternate Optimization Methods

We implemented three additional methods to search for optimal feature sets with respect to feature importance and measurement cost:

1. **Greedy selection**. In the greedy selection version of the algorithm, features are sorted by their importance divided by cost: 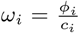. Features are then added to the model one by one, from highest to lowest value of *ω*_*i*_, until no more can be added without exceeding the cost budget. This very simple method works reasonably well.
2. **Recursive feature elimination**. This is inspired by the recursive feature elimination method often used for feature selection in linear models. A model is trained on the full dataset with all features. A measure of feature quality is calculated, and the lowest-quality feature is removed. Another model is trained on the resulting dataset, and the process iterates until only the desired number of features are left. For cost-sensitive prediction, the quality measure is simply importance divided by cost: 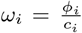, and the iteration stops when model cost is below the budget constraint. This method is valuable for its ties to existing feature selection literature. Because the process is stepwise and feature importance can be recalculated at each iteration, the algorithm can account for feature dependence, where removing one feature changes the importance of other features. Though it performs well it, does not outperform our knapsack method.
3. **Knapsack without retraining**. This is exactly the same as the method described in Method 4, except that after a feature subset is selected, those features are fed into the original model that was trained on all variables. Other features are mean-imputed. This greatly increases training speed but decreases performance, a phenomenon observed in previous work [43].

Performance plots of these alternate methods for the trauma dataset are shown in Supplemental Figure 3.

### Method 7 Base Model Training Details

We used two classes of base models: gradient boosting machines and logistic regression [44, 45]. Hyperparameters were selected using the train/validation splits described in Method 1 with all features then fixed for cost-sensitive learning. The train and validation sets were combined for training after parameters were fixed. We implemented gradient boosting machines using the LightGBM package [40] and used the following parameters:

- Learning rate: 0.01
- Maximum Number of Trees: 1000
- Early Stopping Rounds: 100
- Max Tree Depth: ∈ {1, 2, 4, 8, 10}
- Gamma: 1.0
- Minimum child weight: 10
- Subsampling: ∈ {0.2, 0.5, 0.8, 1.0} We implemented logistic regression models with Scikit-learn [8]and used the following parameters:
- Regularization type: ∈ {L1, L2}
- Regularization strength: ∈ {10^−5^, 10^−4^…10^4^, 10^5^}
- Solver: SAGA

Parameter values not specified above were left at their default values.

### Method 8 Previous Clinical Risk Scores

Several scores have been developed for the specific case of ATC in trauma patients. The COAST score – an additive point-based score using abdominal/pelvic injury, chest decompression, temperature, systolic blood pressure, and entrapment – was one of the earliest [18]. The subsequent PACT score was a six-feature logistic regression involving shock index, age, mechanism of injury, Glasgow Coma Score, and prehospital CPR and intubation [17]. Both models use relatively simple prediction methods and a fixed set of features that limit the range of time budgets in which they can be used. A recent study developed a linear model to predict whether military trauma patients would receive massive transfusion of blood products, with the goal of discovering “concrete and rapidly and easily assessable” predictors. This study did not explicitly account for model cost, but noted that some data, including vital signs, may be difficult to acquire or unavailable in the prehospital setting and developed multiple models with different numbers of features, implying the potential value in this area of models that automatically account for cost [38].

Many risk scores exist to predict mortality of ICU patients. The most popular include the APACHE, APS, SOFA, and qSOFA models [20, 21]. Most of these models take as input a large number of features, while the qSOFA score uses only the Glasgow Coma Score, respiratory rate, and blood pressure at the cost of worse predictive performance. These risk scores all use linear or additive models that aim to either achieve high accuracy with many features, or moderate accuracy with few features. Although mortality prediction in critically ill patients is a topic of great interest in medicine, only a small number of feature sets have been explored. There is no single published model that can make accurate predictions within any feature budget. Finally, although outpatient survival prediction is an important task, we are not aware of a standard clinical tool for this purpose.

### Method 9 Implementation of Existing Clinical Models

We compared to the following clinical models: qSOFA, APS, APACHE IIIa, and APACHE IVa in the ICU dataset, and the PACT score in the trauma dataset. APS, APACHE IIIa, and APACHE IVa were pre-calculated for the ICU dataset. We re-implemented the qSOFA and PACT scores by referring to their respective publications [21, 17]. Notably, the qSOFA score required systolic rather than mean blood pressure as an input variable, so we extracted systolic blood pressure data from the eICU dataset and only gave qSOFA access to this variable. We handled missing data in qSOFA by assuming the corresponding binary variable was false (i.e., the input “respiratory rate greater than 22” was always false if respiratory rate was missing). We handled missing data in PACT with mean imputation. We also found that re-training logistic regression models on the variables in the PACT model substantially improved performance. The final plots show results from the re-trained PACT regression.

In Figure 5, we ranked features in each clinical risk score by their importance. We ranked PACT features by standardized regression coefficient. The qSOFA score assigned equal weight to all features, so the ordering was arbitrary. The APACHE IVa score did not publish standardized regression coefficients, but did publish the reduction in loss from adding each group of features to the model. We used this data to rank features by importance; for each group of features other than the APS score, we divided the group’s loss reduction evenly among the group members to obtain estimates of each feature’s loss reduction. For the APS score features, we divided the total loss reduction for APS features proportional to each feature’s maximum possible univariate contribution to the APS score. Some binary features modulate other features’ univariate contributions; for these features, we assign importance proportional to the difference between the maximum univariate score with the feature on and maximum univariate score with the feature off. Features for which this would result in an importance of 0 are assigned an importance of 1. We divided credit for APS components that depended on multiple features, such as GCS and A-a gradient, evenly among the contributing features. A-a gradient is unique in that it is calculated from PaO2 and several other features but cannot have a nonzero contribution to APS when PaO2 itself has a nonzero univariate contribution; thus, we do not allow it to contribute to PaO2’s importance.

### Method 10 Other Cost-sensitive prediction methods

Cost-sensitive prediction is a topic of growing interest in ML and AI. Established techniques, like the LASSO penalty in regression, encourage models to rely on few of their input features but do not generally incorporate the idea that different features may have different costs [46]. More recent methods have attempted to minimize the feature acquisition cost for each individual prediction while maximizing its accuracy. Methods involve either perturbing an existing model to determine the most important features [47], using decision trees to divide the data into similar groups while penalizing splits that use expensive features [48, 49, 9], or applying reinforcement learning (RL) approaches, which use deep learning to simulate the process of asking for features one at a time and then making a prediction [10, 50, 51, 52]. In this paper, we estimate feature importance using state-of-the art axiomatic methods that guarantee features with a greater effect on the output will be ranked more highly. This can be seen as an improvement on perturbation-based methods and allows CoAI to accurately choose the most important features within a given cost budget [28, 29].

Despite the emergence of methods for low-cost AI, scant research has used these methods to produce risk scores for real clinical problems. Many approaches are evaluated on toy datasets with random or arbitrary feature costs. We know of only one paper that evaluated cost-sensitive methods on a clinical prediction task, which used a Mechanical Turk survey to gather costs from laypeople rather than attempting to synthesize expert opinion [52]. While study was a valuable attempt to reduce the burden of diagnosis for patients, because costs were measured on a 1-10 subjective scale of convenience it is not clear how to add costs together or interpret the resulting total model cost. This results in a model with unclear implications for clinical practice. On the contrary, our work uses costs gathered from expert clinicians in units of minutes or dollars, where total model cost has clear clinical implications.

### Method 11 Extrapolating Nationwide CoAI Deployment

We used precision-recall plots (Supplementary Figure 4) to fix a given level of precision (positive predictive value) for risk scores and compare the resulting recall for each model. Recall can be interpreted as the proportion of positive cases (for trauma, ATC; for ICU, deaths) that are detected by a model. Multiplying recall by the number of positive cases nationwide gives the total number of ATC cases or deaths that would be predicted by the deployed model. Precision, or positive predictive value, is the probability that a predicted positive is a true positive; sufficiently high precision is important to avoid alert fatigue. Selecting a target for precision is difficult and case-dependent, but one study of adverse drug event prediction in electronic health records suggested a target that between 10 and 20 percent of alerts lead to clinical intervention [37, 36]. Based on this result, we set our precision target to 0.2. Because precision is fixed to be equal for all models, a model with a greater recall is clearly better: it can provide advance warning of more cases while maintaining the same ratio of true positives to false positives.

There are roughly 2 million EMS trauma responses in the United States per year [34]; ATC rates vary by region and baseline severity but the base rate in our data would mean 120,000 have ATC [14, 35]. Assuming as above that providers are willing to tolerate 4 false positive alerts for every true positive, deploying the PACT score on all trauma responses would require 30 person-years of data-gathering time and provide early warning of 44,699 ATC cases. Deploying CoAI with the same data-gathering budget would correctly warn of 81,054 cases – an additional 36,355. Deploying CoAI with the EMS provider-preferred budget would reduce the data-gathering requirement to 3 cumulative years, a reduction of 27 total years, while catching 59,301 cases, an additional 14,602 over PACT.

There are over 5 million ICU admissions in the United States per year; the base rate in our data implies roughly 450,000 of those admissions end in death, similar to rates published in previous research [19]. As mentioned in the introduction, the costly APACHE score is not often used in clinical practice, while the relatively inaccurate qSOFA score is easier to use. Thus we compare qSOFA, which uses 3 features, against the 3-feature CoAI model. Here we consider a 3.5:1 false:true ratio, which lines up with one of the few available operating points for qSOFA. Here, qSOFA correctly provides advance warning 182,653 deaths, while CoAI using the same number of features provides advance warning of 389,809 – an increase of 207,156 patients.

### Method 12 CEGB Implementation Details

We implemented Cost-Effective Gradient Boosting using the authors’ code which is integrated into LightGBM [9]. We used the `cegb penalty feature coupled` parameter to pass the per-feature cost vector and `cegb tradeoff` to control the cost-performance tradeoff. The `cegb penalty feature coupled` parameter charges a global cost the first time a feature is used in any tree. While other options exist, such as charging a cost the first time a feature is used for each sample (resulting in different features being measured for different samples), the coupled penalty is the most apt comparison to CoAI, since it selects a low-cost set of features to be measured for all patients. We tuned the `cegb tradeoff` parameter on 101 logarithmically spaced points in [10^−2^, 10^5^].

### Method 13 RL Implementation Details

We implemented the reinforcement learning approach of [10] using the authors’ published code. We used hyperparameter values for most parameters from the largest network included in the example code and set the neural network’s hidden layer size to 128 and the “difficulty” (a multiplier on the number of steps for training, early stopping, etc) to 1000. We tuned the regularization strength with 8 logarithmically spaced points: {10^−7^, 10^−6^…10^0^}. This resulted in fewer points on the cost-performance curve than CoAI or CEGB but was necessary due to the method’s slow runtime.

The reinforcement learning approach was not directly comparable to CoAI because it is allowed to choose different features for each patient. Thus, it may attain a lower cost that is unattainable by a model with a single fixed list of features. However, we could not easily alter this property so we did not change this behavior. In all figures, we use the “average-budget” version of CWCF to provide a generous upper bound on performance, though we note that CWCF is capable of using a hard budget like CoAI at the cost of lower predictive power.

Reinforcement learning suffered in our tests because it makes dichotomous predictions by default, which dramatically reduces its performance in a ranking-based metric like AU-ROC. We attempted to account for this by editing the code to dump the final Q values for each sample at test time. Because Q-values correspond to expected reward for taking an action, we interpret the Q-values for the “classify into a given class” action as a measure of the model’s confidence in predicting that class. Using Q-values as pseudo-probabilities improves classification metrics like AU-ROC but still underperforms, perhaps in part because Q-values in an RL task do not lead to as well-calibrated probabilities as the outputs of a true classification model with a logistic objective. We were also able to run only 17 replicates for the trauma data and 25 for the outpatient data due to long runtime and failure to converge on many runs.

### Method 14 Cost-Performance Curve Comparison

We compared several methods for low-cost AI modeling in Figure 4. We retrained each method on 100 random train-test splits of the dataset. Parameters were fixed on an initial random train/validation/test split and preserved across all subsequent runs. Because CEGB and CWCF may produce models of different costs on each run, we calculated the mean and standard deviation of each model’s cost-performance curves by interpolating them along the same 100 points, linearly spaced between the lowest and highest-cost models over the 100 runs. We used previous-value interpolation so each point was a conservative estimate of performance (i.e., if no 5-minute model existed for a run, the 4-minute model’s performance would be used to interpolate performance at a 5-minute budget).

To calculate statistical significance, we used a two-sided paired-samples T-test, paired by random train-test split, on the mean AU-ROC for each model’s cost-performance curves. The T-test normality assumption is justified by the central limit theorem and the fact that each sample in this test is a *mean* AU-ROC. Because CWCF ran slowly and failed to complete for many runs, comparisons between other methods and CWCF used only paired samples for which CWCF had a complete run available (17 runs for the trauma dataset and 25 for the outpatient dataset). Tables of the resulting p-values are shown in Supplementary Figures 10 and 11.

In the outpatient data, we had to account for the fact that neither CEGB nor CWCF was able to use the information that features came in groups. We post-hoc adjusted CEGB’s model costs by examining the features used by each CEGB model and re-assigning costs based on which *groups* were used. We were not able to post-hoc adjust CWCF’s model costs, because the CWCF package is not aware of feature groups and reports only the model’s total cost, not which features were used. Thus, we calculated an upper bound for CWCF by setting the performance of *all* CWCF models on a given train-test split to the maximum performance of *any* CWCF model on that split. This will never underestimate performance of CWCF at any budget, and will overestimate performance at all but one point.

### Method 15 Grouped Feature Costs

We extended CoAI to handle grouped feature costs, as encountered in the outpatient dataset, by performing the same knapsack optimization but over groups rather than features. Each group *g* has a single cost *c*_*g*_ for acquiring all features in that group. It also has an importance equal to the sum of importances for each feature in the group: *ϕ*_*g*_ = ∑_*i*∈*g*_ *ϕ*_*i*_. We solve the knapsack problem:

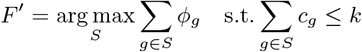

This gives us a set *F* ′ of groups *g* ∈ *F* ′. To identify the features for model training, we simply take the union of all features *i* in all groups in *F* ′: *F* =⋃_*g*_{*i*|*i* ∈ *G*}. The model cost is the sum of group costs: ∑_*g*∈*F*′_ *c*_*g*_.

### Method 16 Binary Search/Tuning Details

We compared the efficiency of model tuning by trying to build optimal models under a particular cost budget with CoAI, CEGB, and reinforcement learning. This was straightforward with CoAI: we entered the desired cost *k* as a model parameter. CoAI trained one full-data model to calculate the *ϕ*_*i*_ values, selected maximum-importance features within that budget, and then trained the final model. It is a less straightforward process in other frameworks to determine how to satisfy a particular cost constraint while guaranteeing that the best possible model within the framework has been found.

In CEGB and reinforcement learning, we had to use blind tuning to achieve the same goal because the relationship between their cost-performance tradeoff parameter *λ* and the actual model cost is unclear and dependent on many factors (magnitude of the loss, scale of the costs, etc). All we know is that as *λ* increases, cost and accuracy should monotonically decrease. Thus, we performed binary search by setting upper and lower bounds on *λ* and a starting value *λ*_0_. For rounds *i* from 0 to *T*, we iteratively train a model with *λ*_*i*_ and check if the model’s cost is below our target *k*. If so, we set 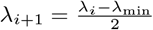. If not, we set 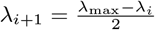. We then continue the iteration.

Values used for this search are:

- *λ*_min_ = 0
- *λ*_max_ = 10^6^
- *λ*_0_ = 1
- *T* = 128

CEGB converged after 28 iterations and was terminated early. CWCF ran for the full 128 iterations.

## Data Availability

Two of our three datasets – the ICU and outpatient datasets – are publicly available. The ICU dataset was published in [39] and is available from the MIT eICU Collaborative Research Database (https://eicu-crd.mit.edu/gettingstarted/overview/) but requires approval before download. The outpatient dataset is a subset of the NHANES I study [22] and was published in its current format in [29]. It is also uploaded to our Github repository along with our code (see below). The trauma dataset is not publicly available due to patient privacy concerns.

## Code Availability

Code implementing CoAI is available at https://github.com/suinleelab/coai. The repository also includes notebooks reproducing the results that do not rely on the trauma dataset, including performance and feature importance for CoAI and existing mortality risk scores on the ICU dataset and comparisons with existing low-cost AI methods on the outpatient dataset.

## Institutional review board statement

The survey data for this study was gathered under an exempt determination from the University of Washington Institutional Review Board (Human Subjects Division, STUDY00006890).

## Footnotes

1 https://github.com/suinleelab/coai

