## Supplementary Figures for "CoAI: Cost-Aware Artificial Intelligence for Health Care"

### 1 Supplementary Figures

| | Measurement Cost (Minutes) |
|---|---|
| Age | 0.28 |
| Agency Level from Scene | 0.00 |
| Agency Mode from Scene | 0.00 |
| Age (Units) | 0.00 |
| Cause of Injury | 0.85 |
| Ethnicity | 0.59 |
| Form from Scene | 0.00 |
| Race | 0.39 |
| Residence State | 0.00 |
| Destination Reason from Scene | 0.00 |
| First Blood Pressure on Scene | 1.99 |
| First Pulse on Scene | 0.81 |
| First Respiration Rate on Scene | 1.09 |
| GCS on Scene (Eyes) | 1.13 |
| GCS on Scene (Motor) | 1.88 |
| GCS on Scene (Verbal) | 1.00 |
| Assisted Respirations on Scene | 0.00 |
| Sex | 0.32 |
| Arrival Date to Scene (Month) | 0.00 |
| Arrival Date to Scene (Day) | 0.00 |
| Arrival Date to Scene (Weekday) | 0.00 |
| Departure Date (Month) | 0.00 |
| Departure Date (Day) | 0.00 |
| Departure Date (Weekday) | 0.00 |
| Injury Date (Month) | 0.00 |
| Injury Date (Day) | 0.00 |
| Injury Date (Weekday) | 0.00 |
| Notification Date to Scene (Month) | 0.00 |
| Notification Date to Scene (Day) | 0.00 |
| Notification Date to Scene (Weekday) | 0.00 |
| Arrival Time to Scene | 0.00 |
| Departure Time from Scene | 0.00 |
| Injury Time | 0.00 |
| Notification Time to Scene | 0.00 |
| Injury ZIP Code (km N of hospital) | 0.00 |
| Injury ZIP Code (km E of hospital) | 0.00 |
| Residence ZIP Code (km N of hospital) | 0.00 |
| Residence ZIP Code (km E of hospital) | 0.00 |
| Intubation (Procedure) | 0.00 |
| Other Splinting/Immobilization (Procedure) | 0.00 |
| IV Placement (Procedure) | 0.00 |
| Cervical Collar (Procedure) | 0.00 |
| Backboard (Procedure) | 0.00 |
| Supplemental Oxygen (Procedure) | 0.00 |
| Pelvic Binder/Sheeting (Procedure) | 0.00 |
| CPR (Procedure) | 0.00 |

Figure 1: Survey Results: Estimated costs for all trauma features

| | Feature Group Cost (Dollars) |
|---|---|
| BUN | 4.39 |
| Age | 0.00 |
| Alkaline Phosphatase | 9.56 |
| CBC w/Diff | 8.63 |
| Calcium | 5.73 |
| Cholesterol | 14.88 |
| Creatinine | 5.69 |
| Height | 0.00 |
| Hemoglobin | 2.63 |
| Physical Activity | 0.00 |
| CBC Auto | 7.18 |
| Potassium | 5.11 |
| Pulse Pressure | 0.00 |
| Red Blood Cells | 3.35 |
| Sedimentation Rate | 3.00 |
| Serum Albumin | 5.50 |
| Serum Protein | 4.07 |
| Sex | 0.00 |
| Sodium | 5.35 |
| Systolic BP | 0.00 |
| Total BIlirubin | 5.57 |
| Uric Acid | 5.02 |
| Urine Albumin | 6.42 |
| Urine Glucose | 4.37 |
| Urinalysis | 2.41 |
| Weight | 0.00 |
| SGOT | 4.83 |

Figure 2: Numeric costs for outpatient features by group.

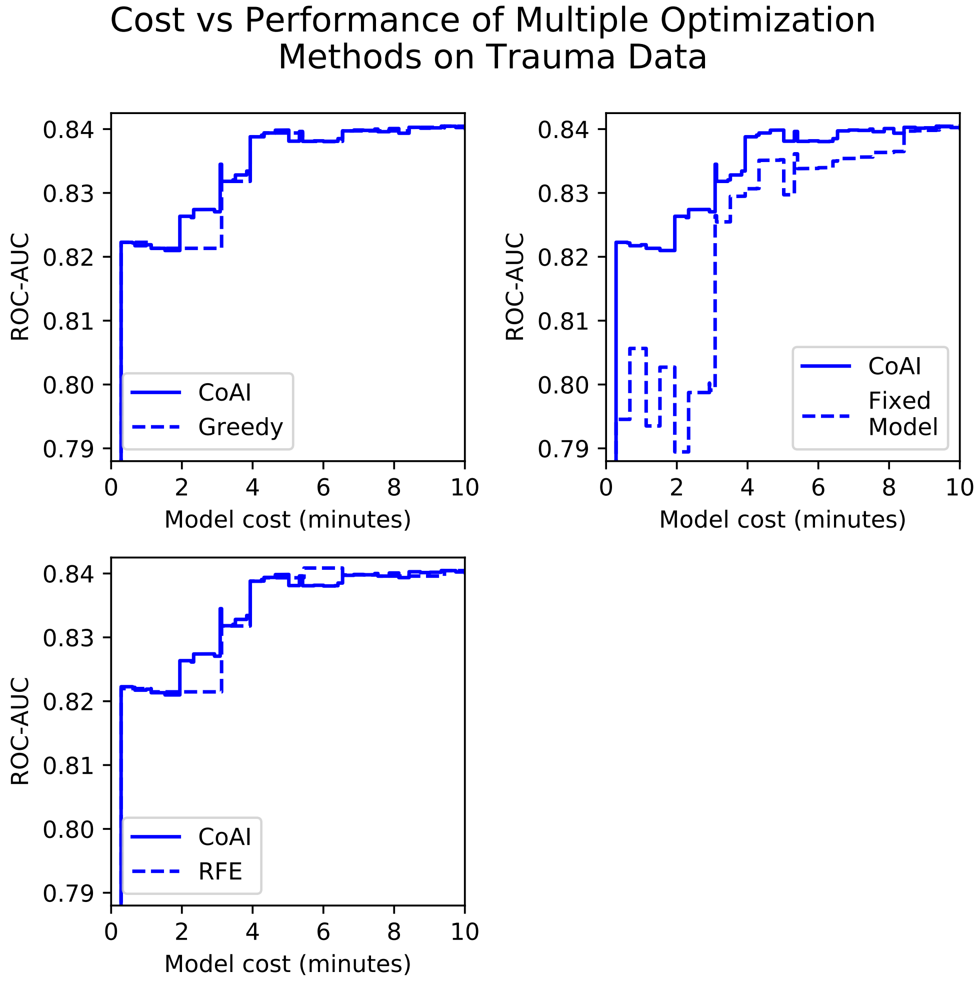


Figure 3: Performance of CoAI variants on the trauma data, including a greedy solution to the knapsack problem where features are added in order of decreasing (importance divided by cost), a method using the same knapsack solver as the maintext but without model retraining, and a recursive feature elimination based method where the feature with lowest (importance divided by cost) is removed from the model, the model is retrained, and the process is repeated until the budget $k$ is satisfied.

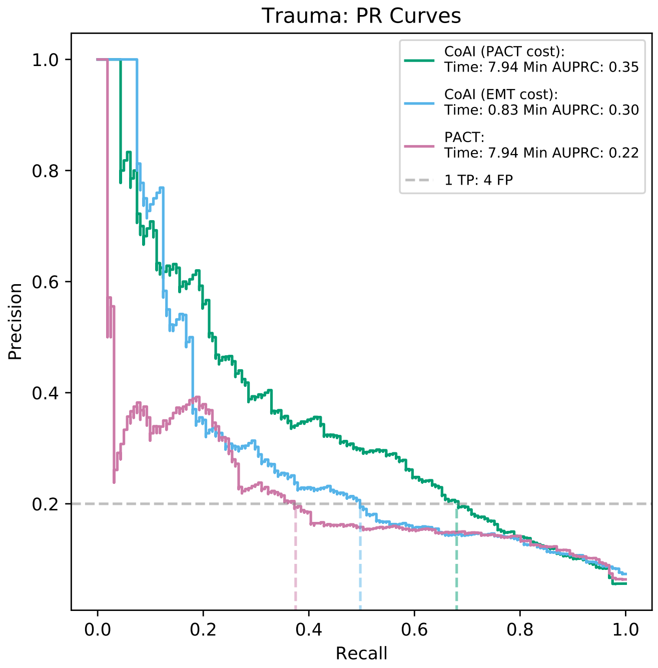


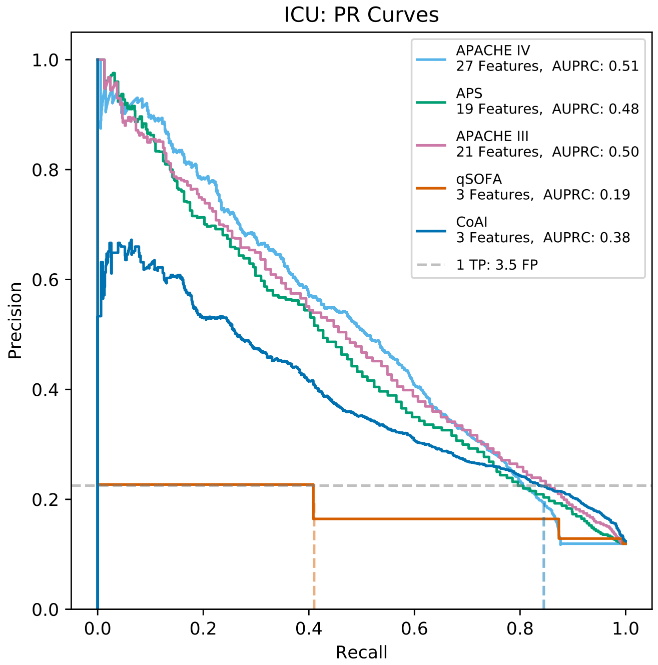


Figure 4: Precision-recall plots for the trauma (top) and ICU (bottom) datasets. Performance curves are shown for CoAI and for the clinical models it was compared against. Horizontal gray lines indicate selected operating points; in the trauma dataset, we chose an operating point with 4 false-positives for every true positive. In the ICU dataset, qSOFA had a limited number of operating points so we compared at a 3.5:1 operating point. Vertical colored lines match model performance at given operating points to the x-axis for easier comparison of recall.

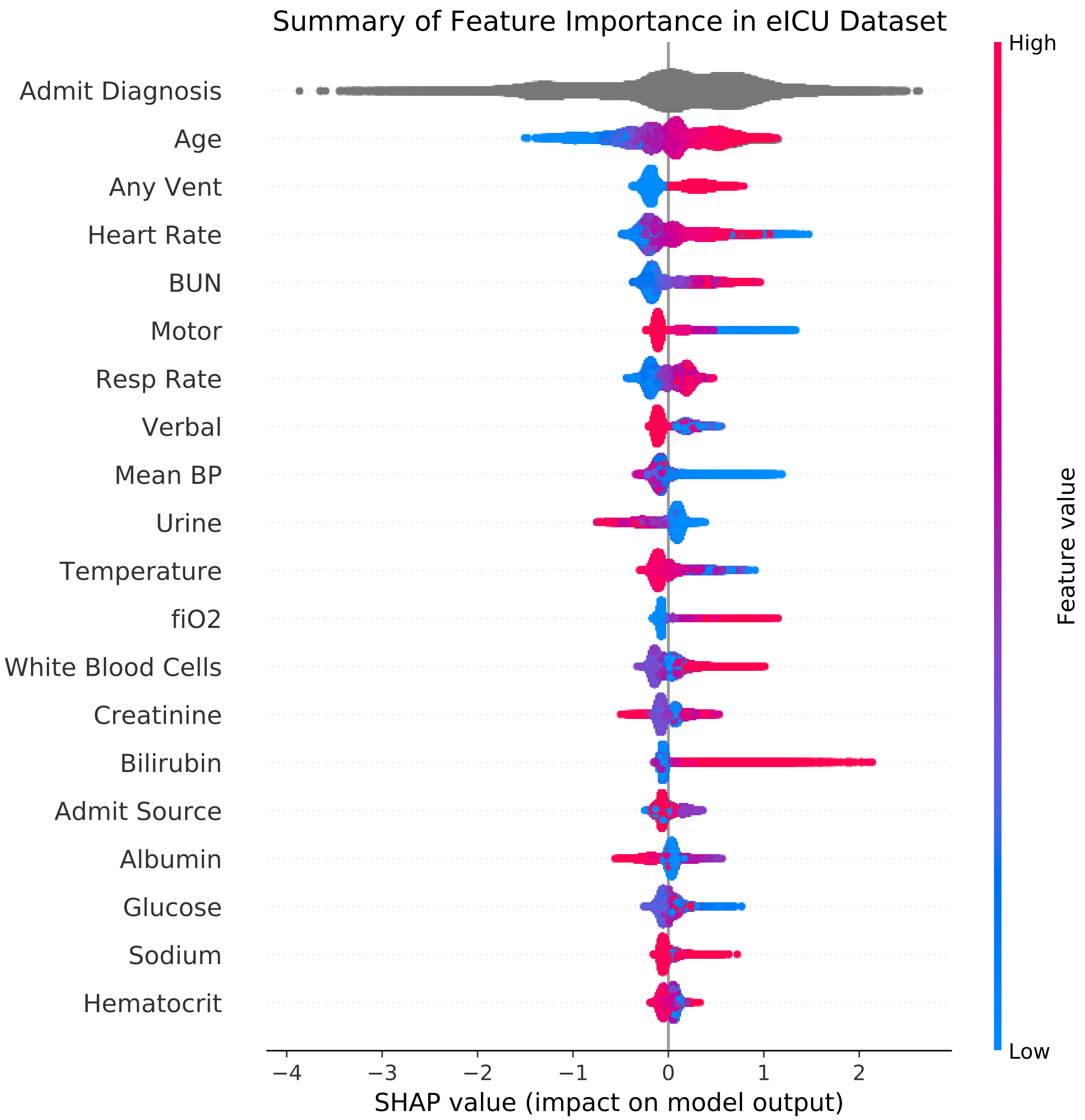


Figure 5: Summary of feature importance for predicting mortality in the eICU dataset.

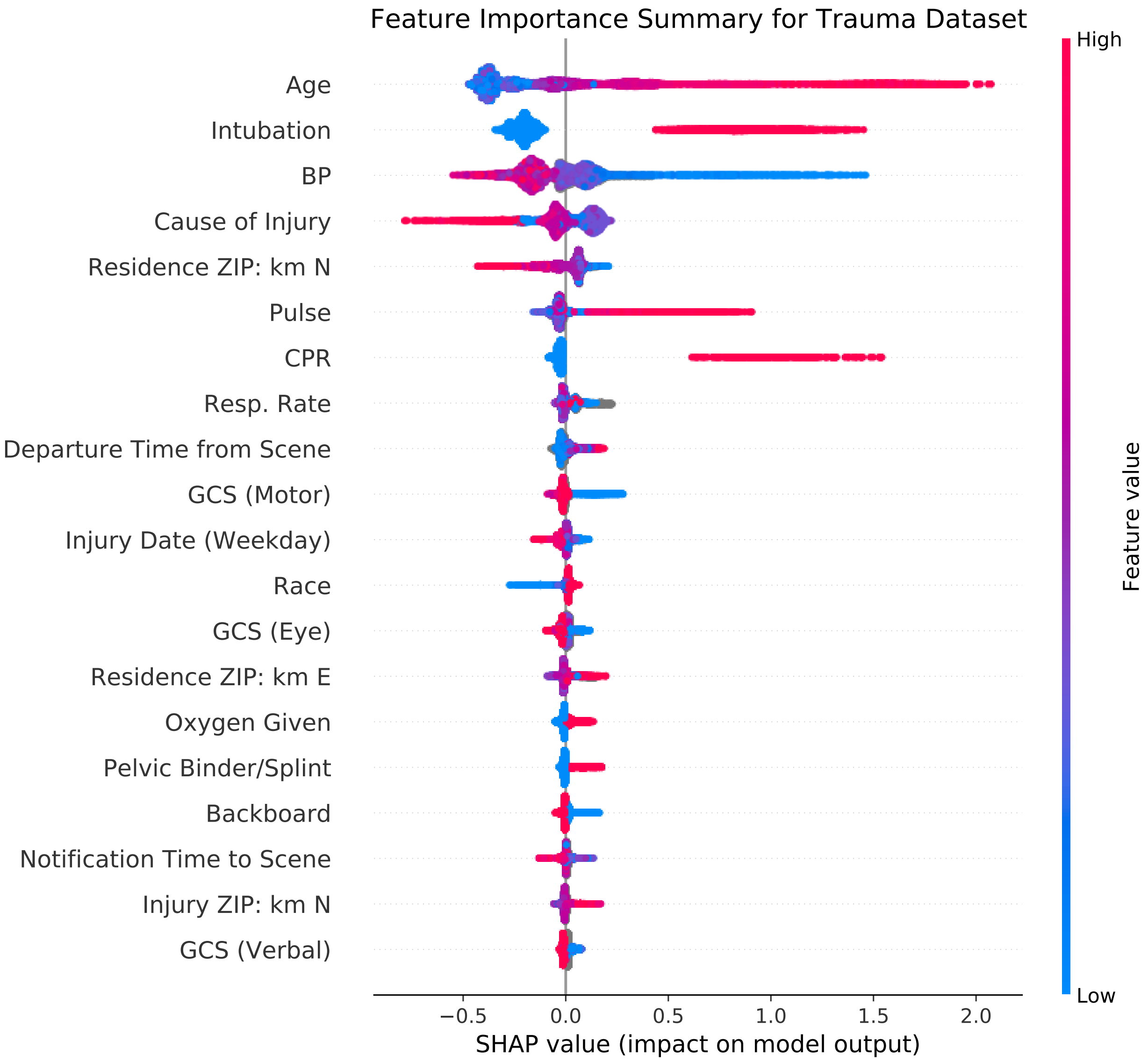


Figure 6: Summary of feature importance for predicting mortality in the trauma dataset.

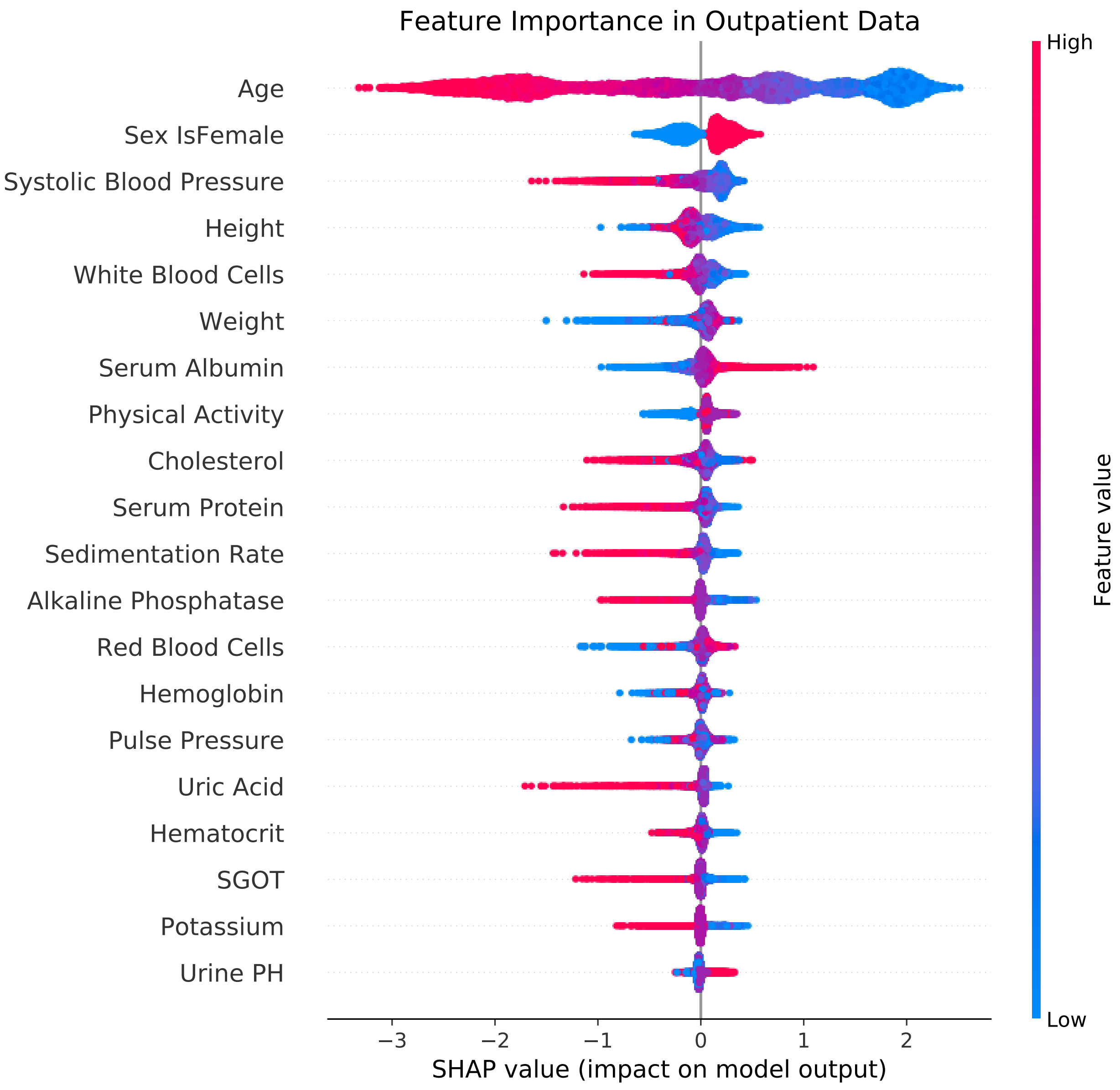


Figure 7: Summary of feature importance for predicting mortality in the outpatient dataset.

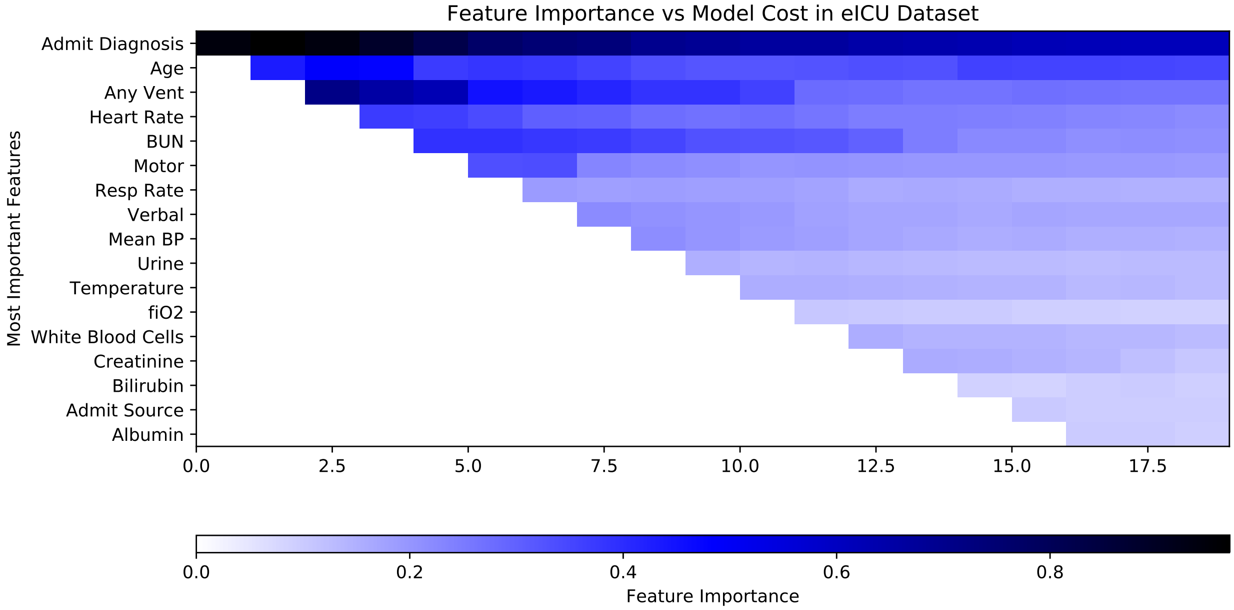


Figure 8: Feature importance heatmap for CoAI on eICU dataset.

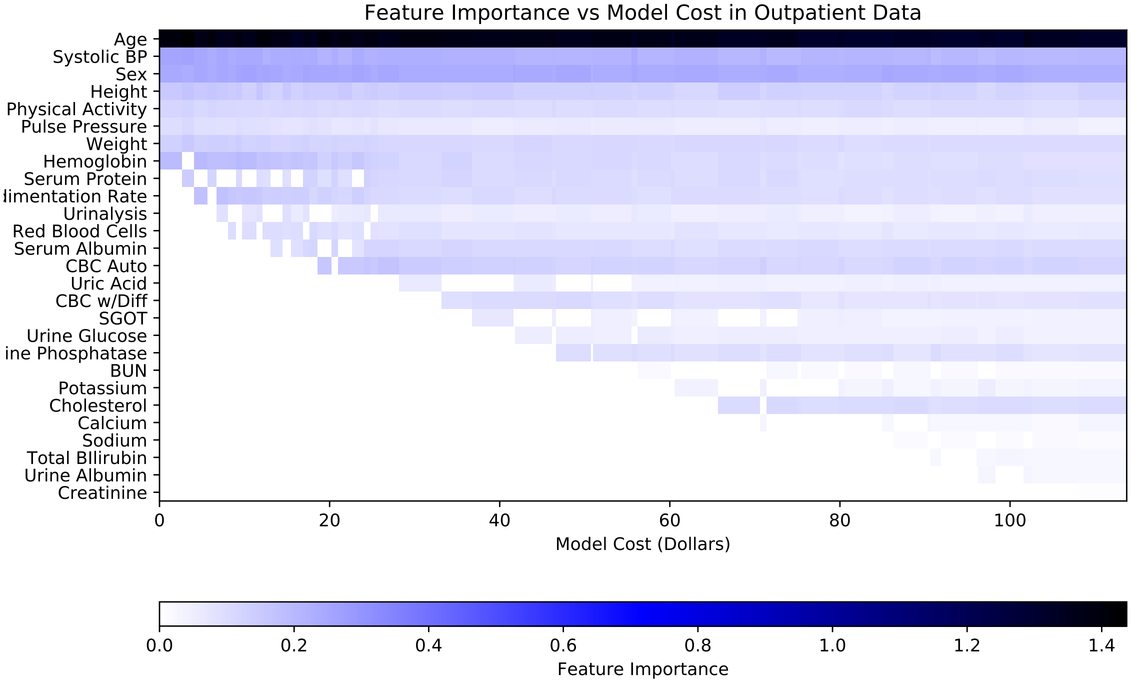


Figure 9: Feature importance heatmap for CoAI on outpatient dataset.

| Method 1 | Method 2 | T-statistic | p-value |
|---|---|---|---|
| CoAI (GBM) | CoAI (Linear) | 27.241980 | 8.569957e-48 |
| CoAI (GBM) | CEGB | 9.970269 | 1.270177e-16 |
| CoAI (GBM) | CWCF | 18.076428 | 4.522429e-12 |
| CoAI (Linear) | CEGB | -26.157262 | 2.943945e-46 |
| CoAI (Linear) | CWCF | 14.619751 | 1.122264e-10 |

Figure 10: Statistical significance of performance differences between all methods on trauma dataset.

| Method 1 | Method 2 | T-statistic | p-value |
|---|---|---|---|
| CoAI (GBM) | CoAI (Linear) | -6.259210 | 9.970512e-09 |
| CoAI (GBM) | CEGB | 5.757907 | 9.590193e-08 |
| CoAI (GBM) | CWCF | 6.094629 | 2.701177e-06 |
| CoAI (Linear) | CEGB | 7.037159 | 2.592167e-10 |
| CoAI (Linear) | CWCF | 7.577163 | 8.139467e-08 |

Figure 11: Statistical significance of performance differences between all methods on outpatient dataset.

### 2 EMS Provider Survey

**Preliminary**

EMS Trauma Scene Response Survey

**Thank you for participating in this survey! In this form, we will ask you for your best estimate of the time or effort required to obtain various types of pre-hospital data about patients that may be useful for diagnosis or risk stratification. The results of this survey will help us build easier-to-use computerized diagnosis and risk-scoring tools.**

Figure 12: Survey form for trauma

Which of the following EMS certifications do you hold?

- ☐ EMT-B
- ☐ AEMT
- ☐ Paramedic
- ☐ Flight paramedic
- ☐ Flight nurse
- ☐ MD

Which EMS agency do you currently work for?

- ☐ Airlift Northwest
- ☐ Other ambulance company
- ☐ Other EMS, not an ambulance company
- ☐ Seattle Fire Medic One
- ☐ King County Medic One
- ☐ Shoreline Medic One
- ☐ Bellevue Medic One
- ☐ Redmond Medic One
- ☐ Falck
- ☐ TriMed
- ☐ AMR

For how many years have you worked in any EMS role?

Figure 13: Survey form for trauma

#### Dispatch Variables

**(Required) How much TIME** does it take to gather the following patient information during a trauma response?

In each case, we are asking for your best estimate, in seconds or minutes, of the total time required to gather the necessary information.

*For reference, information that is provided and known at the time of dispatch should be thought of as requiring zero time.*

PLEASE MOVE THE SLIDER TO RECORD THE TIME REQUIRED FOR EACH ITEM

Figure 14: Survey form for trauma

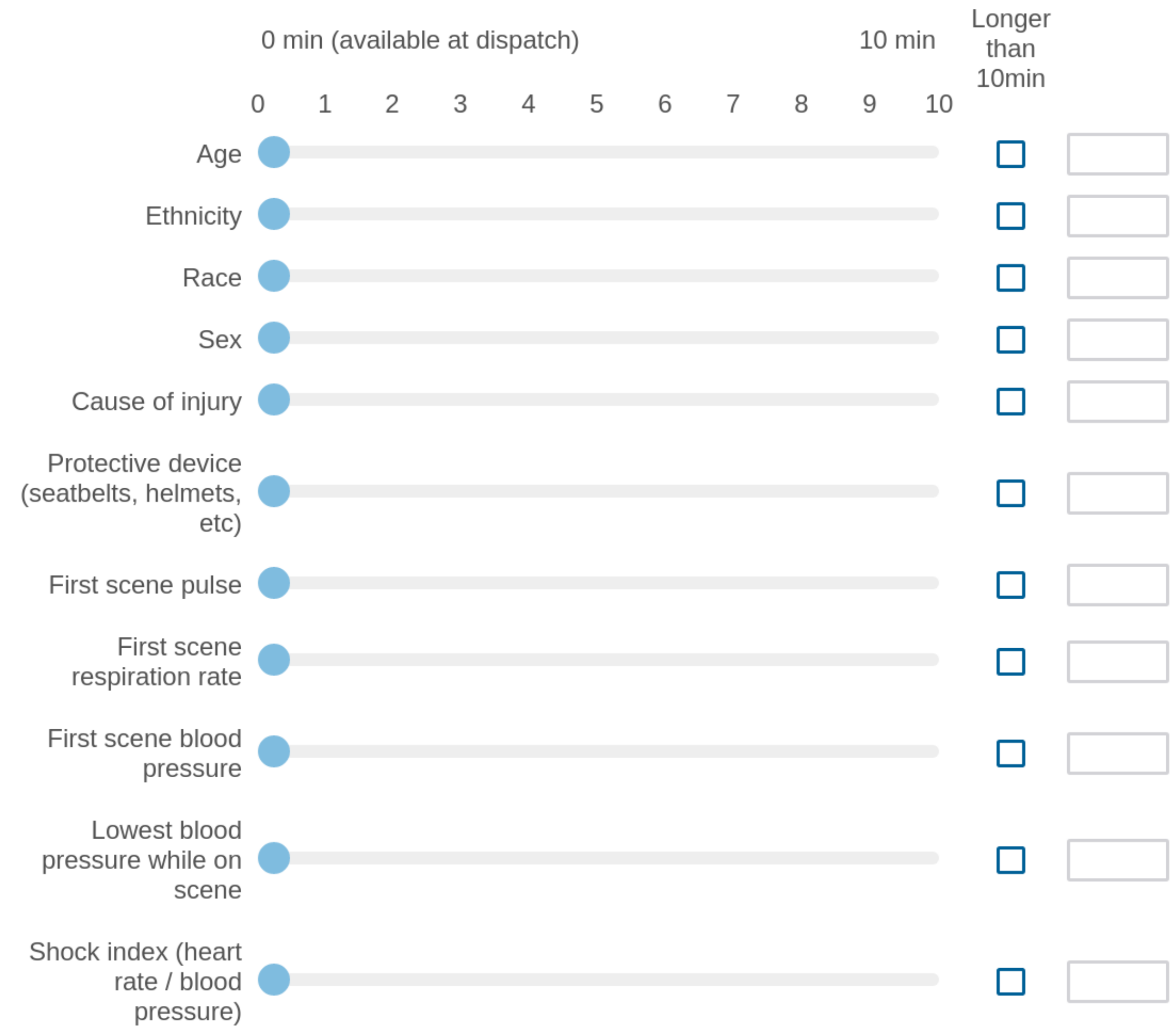


Figure 15: Survey form for trauma

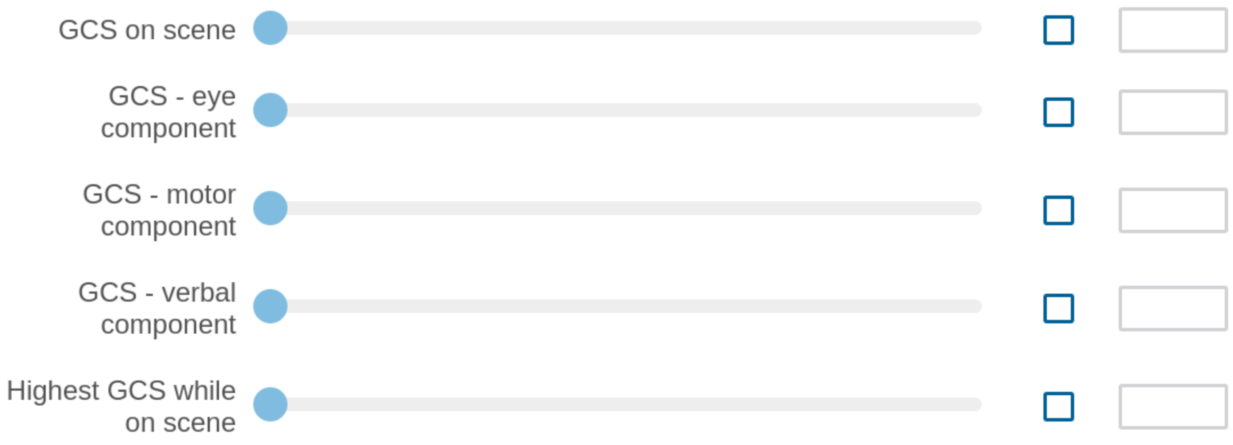


Figure 16: Survey form for trauma

**(Required) HOW DIFFICULT or burdensome** it is to gather the following patient information <u>during a trauma response</u>?

We are asking for your best estimate, as a subjective rating of the amount of effort required, with 1 being the least and 10 being the most, to gather the necessary information.

*For reference, a rating of 1 might be assigned to information that you have been provided at the time of dispatch and requires almost no effort. A rating of 10 might represent information that is very difficult to obtain, due to the need for detailed patient interview or close physical examination or an advanced procedural task.*

PLEASE MOVE THE SLIDER TO RECORD THE LEVEL OF DIFFICULTY FOR EACH ITEM

Figure 17: Survey form for trauma

| | Extremely easy | | | | Moderately Difficult | | | | Extremely Difficult | |
|---|---|---|---|---|---|---|---|---|---|---|
| | 1 | 2 | 3 | 4 | 5 | 6 | 7 | 8 | 9 | 10 |
| Age | | | | | | | | | | |
| Ethnicity | | | | | | | | | | |
| Race | | | | | | | | | | |
| Sex | | | | | | | | | | |
| Cause of injury | | | | | | | | | | |
| Protective device (seatbelts, helmets, etc) | | | | | | | | | | |
| First scene pulse | | | | | | | | | | |
| First scene respiration rate | | | | | | | | | | |
| First scene BP | | | | | | | | | | |
| Lowest BP while on scene | | | | | | | | | | |
| Shock index (heart rate / blood pressure) | | | | | | | | | | |

Figure 18: Survey form for trauma

GCS on scene

GCS - eye component

GCS - motor component

GCS - verbal component

Highest GCS while on scene

Figure 19: Survey form for trauma

**Block 3**

The following variables are ***procedures*** rather than variables to be measured. For each procedure, please provide your best estimate, in seconds or minutes, of the total time required to **recognize that the procedure is necessary and to perform it**.

PLEASE MOVE THE SLIDER TO RECORD THE TIME REQUIRED FOR EACH ITEM

Figure 20: Survey form for trauma

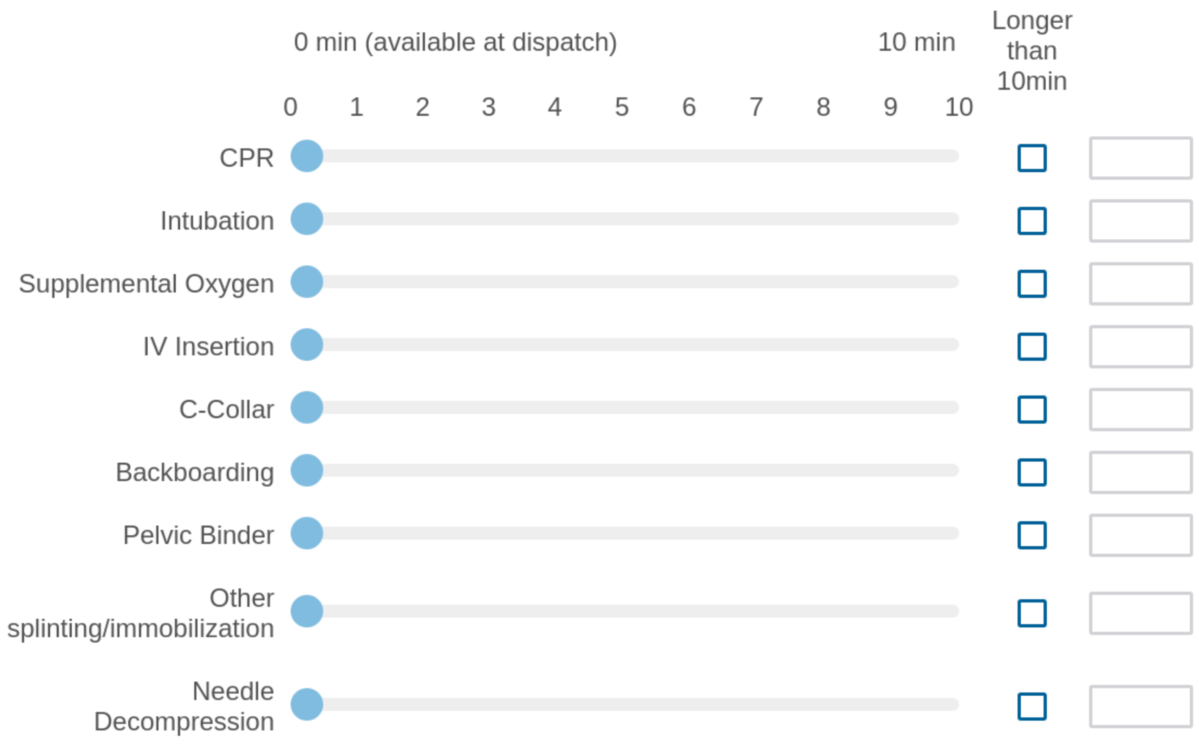


Figure 21: Survey form for trauma

For each procedure, please provide your best estimate of the of the amount of effort required, with 1 being the least and 10 being the most, to **recognize that the procedure is necessary and to perform it.**

PLEASE MOVE THE SLIDER TO RECORD THE EFFORT REQUIRED FOR EACH ITEM

| | Extremely easy | | | | Moderately Difficult | | | | Extremely Difficult | |
|---|---|---|---|---|---|---|---|---|---|---|
| | 1 | 2 | 3 | 4 | 5 | 6 | 7 | 8 | 9 | 10 |
| CPR | | | | | | | | | | |
| Intubation | | | | | | | | | | |
| Supplemental Oxygen | | | | | | | | | | |
| IV Insertion | | | | | | | | | | |
| C-Collar | | | | | | | | | | |
| Backboarding | | | | | | | | | | |
| Pelvic Binder | | | | | | | | | | |
| Other splinting/immobilization | | | | | | | | | | |
| Needle Decompression | | | | | | | | | | |

Figure 22: Survey form for trauma

**Usage Questions**

Do you currently use computerized risk scores during EMS trauma responses?

- ○ Yes
- ○ No

Which computerized risk scores do you use during an EMS response?

Figure 23: Survey form for trauma

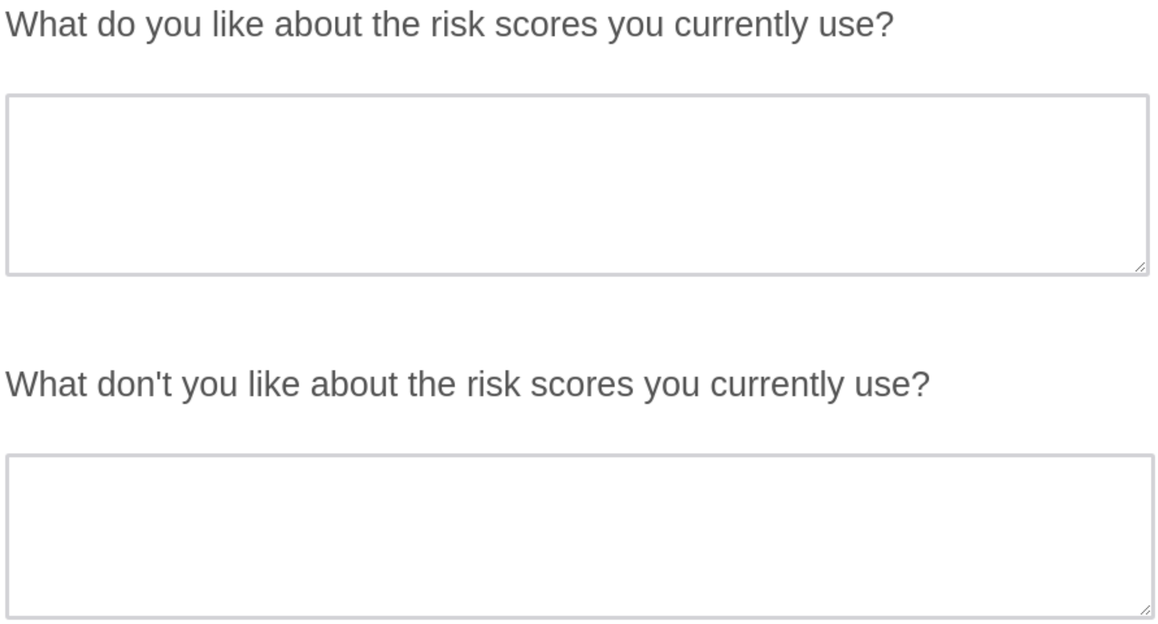
What do you like about the risk scores you currently use?

What don't you like about the risk scores you currently use?

Figure 24: Survey form for trauma

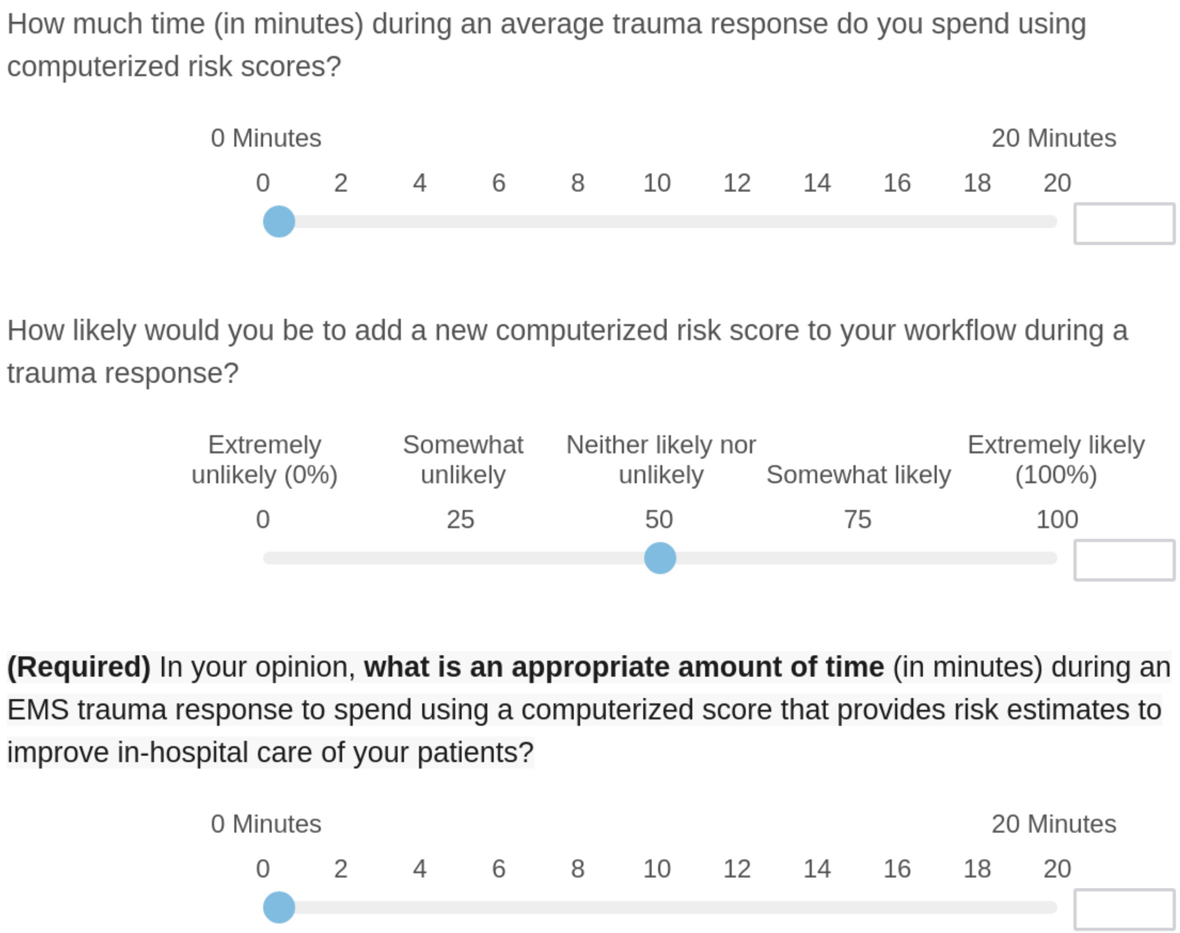

How much time (in minutes) during an average trauma response do you spend using computerized risk scores?

0 Minutes | 20 Minutes

0 2 4 6 8 10 12 14 16 18 20

How likely would you be to add a new computerized risk score to your workflow during a trauma response?

Extremely unlikely (0%) | Somewhat unlikely | Neither likely nor unlikely | Somewhat likely | Extremely likely (100%)

0 25 50 75 100

**(Required)** In your opinion, **what is an appropriate amount of time** (in minutes) during an EMS trauma response to spend using a computerized score that provides risk estimates to improve in-hospital care of your patients?

0 Minutes | 20 Minutes

0 2 4 6 8 10 12 14 16 18 20

Figure 25: Survey form for trauma

What factors would make you more or less likely to use a computerized risk score during an EMS trauma response?

Is there anything else you'd like to share?

Figure 26: Survey form for trauma

### 3 EMS Provider Survey Results

| # | Field | Choice Count | |
|---|---|---|---|
| 1 | EMT-B | 8.70% | 2 |
| 2 | AEMT | 0.00% | 0 |
| 3 | Paramedic | 69.57% | 16 |
| 4 | Flight paramedic | 0.00% | 0 |
| 5 | Flight nurse | 17.39% | 4 |
| 6 | MD | 4.35% | 1 |
| | | | 23 |

Showing rows 1 - 7 of 7

Figure 27: Survey results - certifications held by respondents

| # | Field | Choice Count | |
|---|---|---|---|
| 1 | Airlift Northwest | 22.73% | 5 |
| 2 | Other ambulance company | 4.55% | 1 |
| 3 | Other EMS, not an ambulance company | 0.00% | 0 |
| 4 | Seattle Fire Medic One | 72.73% | 16 |
| 5 | King County Medic One | 0.00% | 0 |
| 6 | Shoreline Medic One | 0.00% | 0 |
| 7 | Bellevue Medic One | 0.00% | 0 |
| 8 | Redmond Medic One | 0.00% | 0 |
| 9 | Falck | 0.00% | 0 |
| 10 | TriMed | 0.00% | 0 |
| 11 | AMR | 0.00% | 0 |
| | | | 22 |

Showing rows 1 - 12 of 12

Figure 28: Survey results - Agencies where respondents were employed.

| # | Field | Minimum | Maximum | Mean | Std Deviation | Variance | Count |
|---|---|---|---|---|---|---|---|
| 1 | For how many years have you worked in any EMS role? | 3.00 | 37.50 | 20.43 | 8.96 | 80.28 | 22 |

Figure 29: Survey results - years of prior EMS experience.

| # | Field | Minimum | Maximum | Mean | Std Deviation | Variance | Count |
|---|---|---|---|---|---|---|---|
| 1 | Age | 0.00 | 1.00 | 0.25 | 0.34 | 0.12 | 13 |
| 2 | Ethnicity | 0.00 | 3.00 | 0.55 | 0.86 | 0.74 | 10 |
| 3 | Race | 0.00 | 1.00 | 0.46 | 0.38 | 0.14 | 10 |
| 4 | Sex | 0.00 | 1.00 | 0.29 | 0.27 | 0.07 | 13 |
| 5 | Cause of injury | 0.00 | 3.00 | 0.81 | 0.83 | 0.70 | 13 |
| 6 | Protective device (seatbelts, helmets, etc) | 0.10 | 4.00 | 1.45 | 1.37 | 1.88 | 12 |
| 7 | First scene pulse | 0.10 | 2.00 | 0.76 | 0.61 | 0.37 | 12 |
| 8 | First scene respiration rate | 0.10 | 3.80 | 1.03 | 0.97 | 0.94 | 12 |
| 9 | First scene blood pressure | 0.10 | 4.20 | 1.87 | 1.18 | 1.40 | 12 |
| 10 | Lowest blood pressure while on scene | 0.00 | 7.00 | 2.13 | 1.91 | 3.65 | 12 |
| 11 | Shock index (heart rate / blood pressure) | 0.20 | 10.00 | 2.38 | 2.50 | 6.26 | 12 |
| 12 | GCS on scene | 0.30 | 6.40 | 1.76 | 1.72 | 2.97 | 12 |
| 13 | GCS - eye component | 0.10 | 6.00 | 1.04 | 1.62 | 2.62 | 12 |
| 14 | GCS - motor component | 0.10 | 7.20 | 1.74 | 2.30 | 5.31 | 12 |
| 15 | GCS - verbal component | 0.10 | 6.10 | 0.93 | 1.60 | 2.56 | 12 |
| 16 | Highest GCS while on scene | 0.20 | 10.00 | 2.97 | 3.13 | 9.78 | 12 |

Figure 30: Survey results - Estimated time costs of gathering each feature

| # | Field | Minimum | Maximum | Mean | Std Deviation | Variance | Count |
|---|---|---|---|---|---|---|---|
| 1 | Age | 1.00 | 5.00 | 1.69 | 1.07 | 1.14 | 13 |
| 2 | Ethnicity | 1.00 | 10.00 | 3.92 | 3.17 | 10.07 | 13 |
| 3 | Race | 1.00 | 10.00 | 3.38 | 3.18 | 10.08 | 13 |
| 4 | Sex | 1.00 | 5.00 | 1.54 | 1.08 | 1.17 | 13 |
| 5 | Cause of injury | 1.00 | 4.00 | 2.54 | 0.84 | 0.71 | 13 |
| 6 | Protective device (seatbelts, helmets, etc) | 1.00 | 4.00 | 2.46 | 0.93 | 0.86 | 13 |
| 7 | First scene pulse | 1.00 | 3.00 | 1.77 | 0.58 | 0.33 | 13 |
| 8 | First scene respiration rate | 1.00 | 4.00 | 2.31 | 1.26 | 1.60 | 13 |
| 9 | First scene BP | 1.00 | 5.00 | 2.54 | 1.08 | 1.17 | 13 |
| 10 | Lowest BP while on scene | 1.00 | 10.00 | 3.08 | 2.27 | 5.15 | 13 |
| 11 | Shock index (heart rate / blood pressure) | 1.00 | 10.00 | 3.31 | 2.20 | 4.83 | 13 |
| 12 | GCS on scene | 1.00 | 10.00 | 3.15 | 2.44 | 5.98 | 13 |
| 13 | GCS - eye component | 1.00 | 3.00 | 1.77 | 0.70 | 0.49 | 13 |
| 14 | GCS - motor component | 1.00 | 4.00 | 2.08 | 0.83 | 0.69 | 13 |
| 15 | GCS - verbal component | 1.00 | 3.00 | 1.92 | 0.73 | 0.53 | 13 |
| 16 | Highest GCS while on scene | 1.00 | 10.00 | 2.85 | 2.44 | 5.98 | 13 |

Figure 31: Survey results - Estimated effort costs of gathering each feature

| # | Field | Minimum | Maximum | Mean | Std Deviation | Variance |
|---|---|---|---|---|---|---|
| 1 | CPR | 0.00 | 0.80 | 0.33 | 0.21 | 0.05 |
| 2 | Intubation | 1.00 | 9.90 | 4.46 | 2.86 | 8.17 |
| 3 | Supplemental Oxygen | 0.30 | 3.00 | 1.20 | 0.81 | 0.66 |
| 4 | IV Insertion | 0.50 | 5.30 | 2.63 | 1.55 | 2.41 |
| 5 | C-Collar | 0.50 | 4.00 | 1.81 | 1.03 | 1.05 |
| 6 | Backboarding | 0.50 | 6.70 | 3.08 | 2.00 | 4.00 |
| 7 | Pelvic Binder | 0.80 | 6.60 | 2.96 | 1.90 | 3.62 |
| 8 | Other splinting/immobilization | 0.80 | 7.50 | 3.79 | 1.71 | 2.92 |
| 9 | Needle Decompression | 0.40 | 8.70 | 3.15 | 2.38 | 5.69 |

Figure 32: Survey results - Time costs of performing and recording each of the following procedures.

| # | Field | Minimum | Maximum | Mean | Std Deviation | Variance |
|---|---|---|---|---|---|---|
| 1 | CPR | 1.00 | 8.00 | 1.83 | 1.91 | 3.64 |
| 2 | Intubation | 1.00 | 8.00 | 4.08 | 2.22 | 4.91 |
| 3 | Supplemental Oxygen | 1.00 | 6.00 | 1.58 | 1.44 | 2.08 |
| 4 | IV Insertion | 1.00 | 5.00 | 2.42 | 1.19 | 1.41 |
| 5 | C-Collar | 1.00 | 3.00 | 1.50 | 0.65 | 0.42 |
| 6 | Backboarding | 1.00 | 3.00 | 1.83 | 0.90 | 0.81 |
| 7 | Pelvic Binder | 1.00 | 4.00 | 2.33 | 1.03 | 1.06 |
| 8 | Other splinting/immobilization | 1.00 | 7.00 | 2.58 | 1.55 | 2.41 |
| 9 | Needle Decompression | 1.00 | 7.00 | 3.50 | 1.55 | 2.42 |

Figure 33: Survey results - Effort costs of performing and recording each of the following procedures.

| # | Field | Minimum | Maximum | Mean | Std Deviation | Variance | Count |
|---|---|---|---|---|---|---|---|
| 1 | Do you currently use computerized risk scores during EMS trauma responses? | 2.00 | 2.00 | 2.00 | 0.00 | 0.00 | 12 |

| # | Field | Choice Count | |
|---|---|---|---|
| 1 | Yes | 0.00% | 0 |
| 2 | No | 100.00% | 12 |
| | | | 12 |

Showing rows 1 - 3 of 3

Figure 34: Survey results - Percentage of respondents who had used a computerized risk score (0 percent).

| # | Field | Minimum | Maximum | Mean | Std Deviation | Variance | Count |
|---|---|---|---|---|---|---|---|
| 1 | 1 | 0.00 | 0.00 | 0.00 | 0.00 | 0.00 | 0 |

Figure 35: Survey results - Amount of time respondents had spent using computerized risk scores in the field (0 minutes).

| # | Field | Minimum | Maximum | Mean | Std Deviation | Variance | Count |
|---|---|---|---|---|---|---|---|
| 1 | How likely would you be to add a new computerized risk score to your workflow during a trauma response? | 0.00 | 75.00 | 34.08 | 24.61 | 605.41 | 12 |

Figure 36: Survey results - Likelihood respondents would be willing to add a new risk score to their workflow.

| # | Field | Minimum | Maximum | Mean | Std Deviation | Variance | Count |
|---|---|---|---|---|---|---|---|
| 1 | 1 | 0.00 | 2.00 | 0.83 | 0.80 | 0.64 | 12 |

Figure 37: Survey results - Amount of time in minutes respondents were willing to spend gathering data for a computerized risk score.
